# Early Antibody Responses Associated with Survival in COVID19 Patients

**DOI:** 10.1101/2021.02.21.21252168

**Authors:** Zhao-Hua Zhou, Sai Dharmarajan, Mari Lehtimaki, Susan L. Kirshner, Steven Kozlowski

## Abstract

Neutralizing antibodies to the SARS CoV-2 spike proteins have been issued Emergency Use Authorizations and are a likely mechanism of vaccines to prevent COVID-19. However, benefit of treatment with monoclonal antibodies has only been observed in clinical trials in outpatients with mild to moderate COVID-19 but not in patients who are hospitalized and/or have advanced disease. To address this observation, we evaluated the timing of anti SARS-CoV-2 antibody production in hospitalized patients with the use of a highly sensitive multiplexed bead-based immunoassay allowing for early detection of antibodies to SARS-CoV-2. We found that significantly lower levels of antibodies to the SARS-CoV-2 spike protein in the first week after symptom onset were associated with patients who expired as compared to patients who were discharged. We also developed a model, based on antibody level trajectory, to predict COVID 19 outcome that is compatible with greater antibody benefit earlier in COVID 19 disease.

**Author Summary:** We evaluated antibodies to SARS-CoV-2 over time in patients that were hospitalized with COVID 19. Early detection of Anti-SARS-CoV-2 antibodies was associated with survival in patients hospitalized with COVID 19. Early antibody levels predicted outcome in our study. This result is consistent with the benefit of therapeutic antibodies early in the course of COVID 19 disease. With additional study, early antibody levels may be helpful in deciding on appropriate therapies.

## Introduction

SARS-COV-2 has led to more than 100 million cases of COVID-19 globally with high morbidity and mortality. There were over 485,000 deaths in the United States as of February 2021 (1). Neutralizing antibodies to the SARS COV-2 spike protein are candidates for therapeutics (2). There have been three Emergency Use Authorizations issued for such antibodies and combinations (3-5), with many others in the development pipeline. Induction of neutralizing antibodies in immunized people is also likely to be a mechanism of vaccines to prevent COVID-19 (6, 7).

COVID-19 has an inflammatory phase associated with Acute Respiratory Distress Syndrome and severe disease (8). This phase has macrophage and monocyte activation, cytokine release, may involve viral transmission that is not dependent on the ACE2 viral receptor (9) and may be enhanced by binding of SARS-CoV-2 specific antibodies to cells via Fc receptors (10). A host with activated immune and endothelial cells may also be more sensitive to antibody-virus immune complex-associated inflammation.

Some studies evaluating neutralizing antibody treatment in patients who are hospitalized and/or have advanced disease have been stopped due to lack of benefit (11, 12). Current EUAs for neutralizing antibody are limited to outpatients with mild to moderate disease who are at high risk for progressing to severe disease (3-5). In contrast to findings of clinical benefit in outpatients early in disease progression with mild to moderate COVID-19, some studies in hospitalized patients have correlated high levels of neutralizing antibody and “early” seroconversion (8-16 days after onset as defined by these studies) with severity of disease for both SARS-Cov-1 (13) and SARS-CoV-2 (14).

To address this apparent gap, we evaluated the timing of anti SARS-CoV-2 antibody generation in patients who survived and in patients who expired with the use of a highly sensitive multiplexed bead-based immunoassay method (15) allowing for earlier detection (within days of symptom onset) of antibody to SARS-CoV-2. We also developed a model that was predictive of outcome based on the trajectory of antibody levels over time.

These results are compatible with a model of COVID-19 disease with an early viral replication phase, wherein antibody may be more beneficial, and a late inflammatory pathology phase, where there is no current evidence of benefit from virus specific antibody. This disease model is supported by convalescent plasma therapy that suggests better outcomes with early treatment (16, 17) and no benefit with late treatment (18).

## Results

Demographics of the patients in the study are provided in subsets by outcome in Table 1. The numbers are small but a notable difference in outcome by sex is observed. There were 4 to 16 collections per patient over a range of 0 to 42 days post-symptom onset. Antibody levels were plotted by time based on days post-symptom onset in the expired and discharged patients for the SARS-CoV-2 antigens in Figure 1. The trajectories of antibody levels appear to be different between hospitalized patients who expired and those who were discharged. Specifically, patients in the expired group seem to have lower antibody levels in the first week to 12 days after onset, after which values in the groups appear to converge.

**Table 1.** Demographics data by patient outcome.

|  | Discharged | Expired | Total |
| --- | --- | --- | --- |
| **Patients** | **22** | **11** | **33** |
| **Age <sup>1</sup>** |  |  |  |
| 30-51 | 5 | 2 | 7 |
| 52-73 | 12 | 6 | 18 |
| 74-95 | 5 | 3 | 8 |
| **Sex** |  |  |  |
| female | 10 | 1 | 11 |
| male | 12 | 10 | 22 |
| **Days from onset to hospitalization <sup>1</sup>** |  |  |  |
| 0-4 | 14 | 8 | 22 |
| 5-9 | 6 | 3 | 9 |
| 10-14 | 2 | 0 | 2 |
| **Treatments <sup>2</sup>** |  |  |  |
| Ventilator/Intubation | 12 | 8 | 20 |
| Convalescent Sera/Plasma | 1 | 3 | 4 |
| Statin | 2 | 2 | 4 |
| **Underlying conditions <sup>3</sup>** |  |  |  |
| Diabetes | 9 | 4 | 13 |
| Hypertension | 10 | 6 | 16 |
| Kidney disease | 3 | 4 | 7 |
| Obese or morbidly obese | 5 | 1 | 6 |
| **Antibody sampling** |  |  |  |
| Samples/patient (Median) | 6 | 6 | 6 |
| Samples/patient (Range) | 4-16 | 4-13 | 4-16 |
| Days post-onset (Range) | 0-42 | 0-30 | 0-42 |

<sup>1</sup> Age and days from onset to hospitalization are divided in three even groups spanning the range of patient data.
<sup>2</sup> Treatments shown were chosen based on potential effect on antibody titer and number of patients under treatment.
<sup>3</sup> Underlying conditions shown were chosen based on prevalence in the patients in the study with 5 or more patients with each condition.

**Figure 1.**
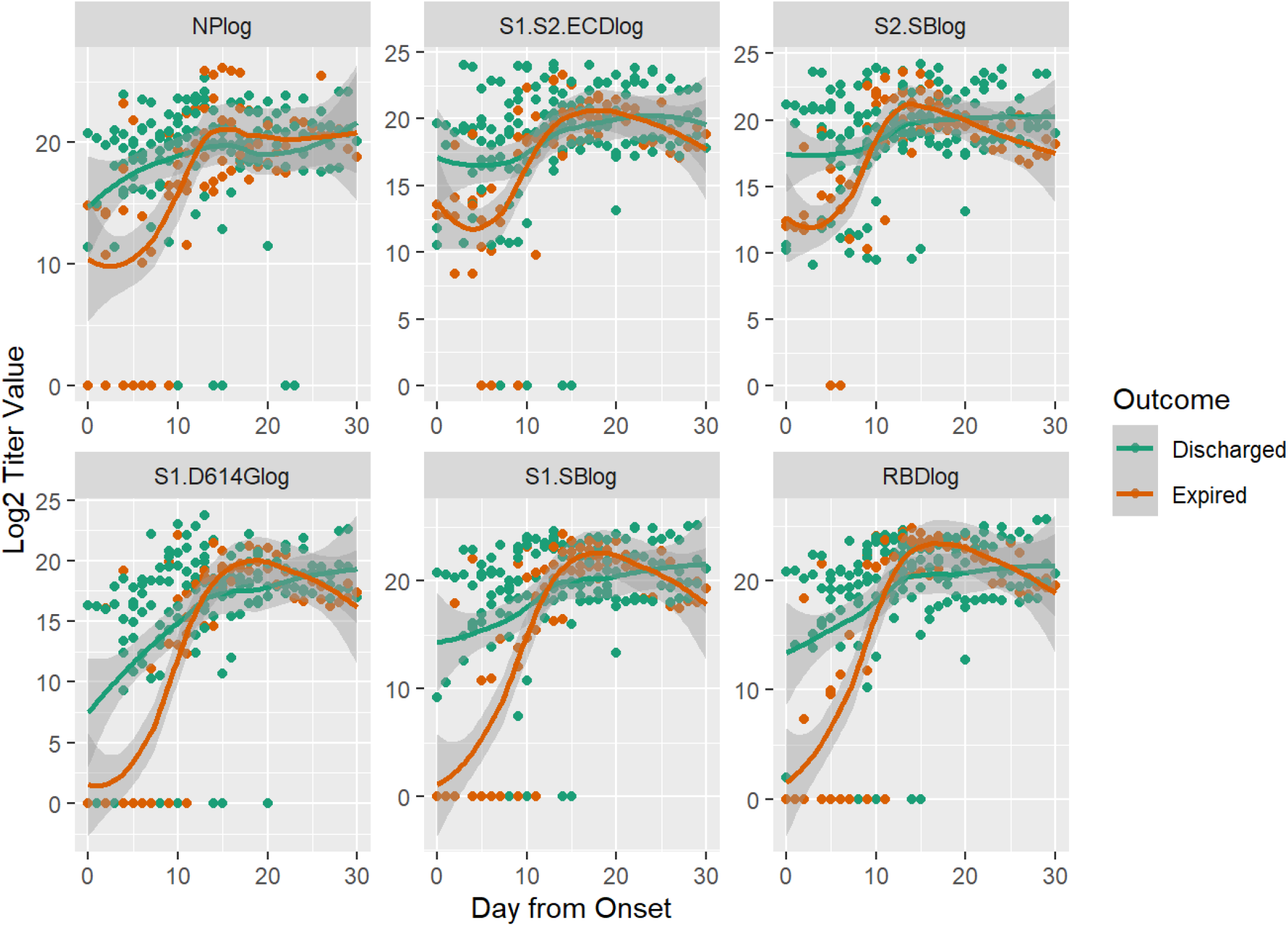
Anti-SARS CoV2 IgG Antibody Kinetics After Symptom Onset. Antibody titers at different days after onset were measured by the multiplexed beads array as described in Methods. Displayed are specific IgG antibody titer values (log transformed) in expired (orange) and discharged (green) groups of patients targeting the nucleocapsid protein (NP) and five spike protein components, i.e., S1+S2 ECD, S2, S1 (D614G), S1 and RBD. Dots are titer values for each patient at the corresponding day from onset; lines are smoothed regression fit to the observed data with 95% confidence interval bands.

Given the amount of missing data in daily antibody levels, we tested for differences across expired and discharged groups using antibody levels at week one, week two and after week two. For patients with more than one sample within a week, we used the median of the available antibody levels. Figure 2 displays the distribution of antibody levels to different antigen targets at week one along with results from a t-test comparing the means of the distributions. Levels of antibodies that bind to all the spike protein antigens and the receptor binding domain were significantly lower at week one for expired patients. The anti-NP antibody levels were also lower at week one for expired patients. Using an alternative maximum signal calculation (inverse dilution x MFI), led to the same findings (Supplementary Materials). No significant differences were seen for antibody levels in week 2; However, after week 2 there was an increased anti-RBD level for expired patients in the primary analysis (Supplementary Materials).

**Figure 2.**
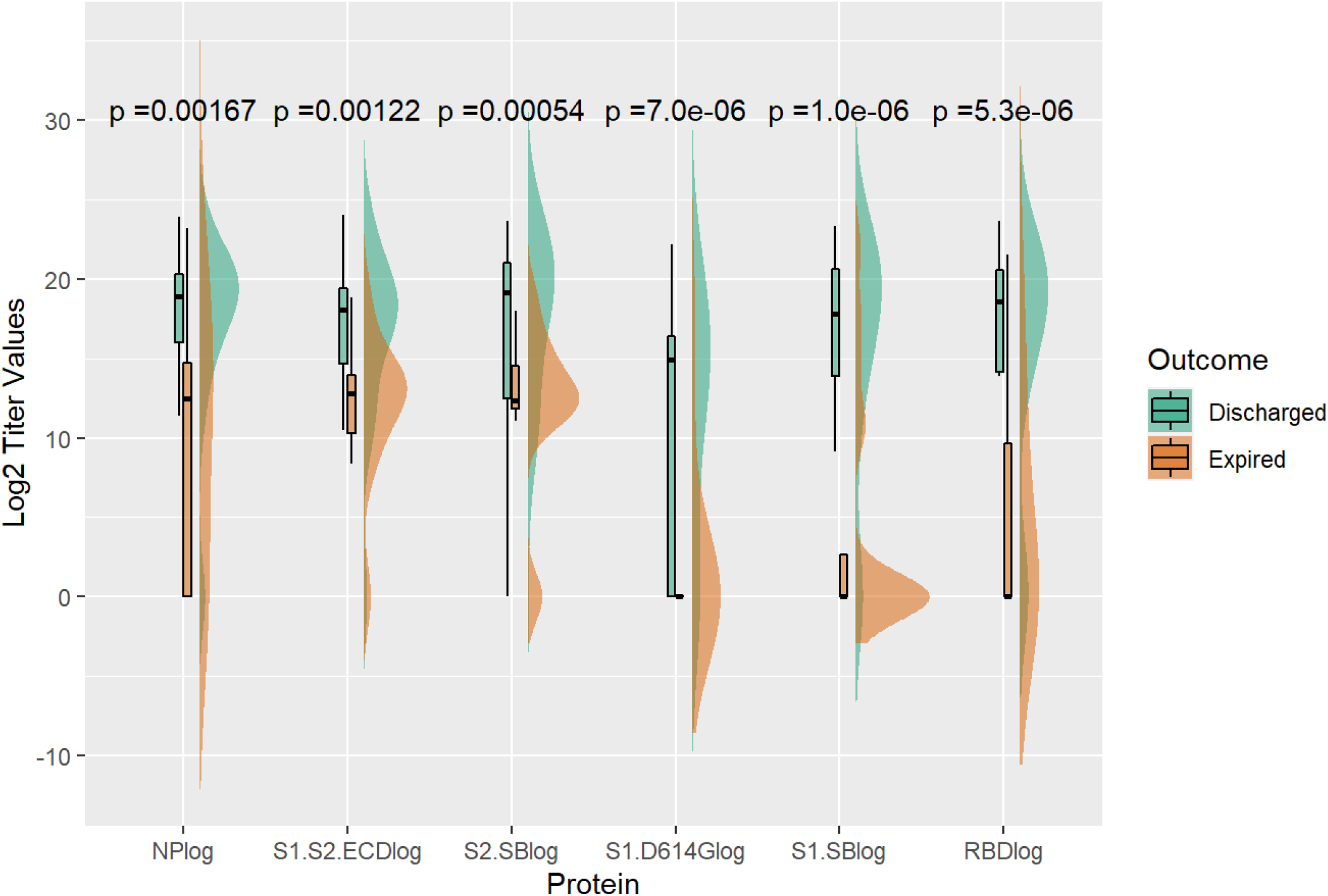
Comparison of Week1 (0 to 7 days) After Onset Anti-SARS COV2 IgG Antibody Titers Between Discharged and Expired Groups. Density and box plots show distribution of specific IgG titer values (log transformed) at week 1 after onset targeting SARS CoV2 NP and five spike protein components, S1+S2 ECD, S2, S1 (D614G), S1 and RBD. The expired (orange) and discharged (green) groups were shown with t-test comparison p-values.

We also fit a linear mixed model (Model 1, Supplementary Materials), accounting for patient-level correlation, to the data to model the antibody response using time, expired / discharged status, and the interaction between time and expired / discharged status as predictors. This model yielded similar results as the t-test in Figure 2. The mean antibody levels, at week one, were estimated to be significantly lower in the expired group than in the discharged group (Table 2). The mean increase from week one was also found to be significantly higher in the expired group at week two for both S1 and S2 spike proteins and at after week 2 for S1 spike protein (Table 2). These findings suggest that later increases in antibody response are unlikely to mitigate the negative effects of an initial slow response.

**Table 2.** Estimates from Linear Mixed Model (Model 1) of Anti-S1 and Anti-S2 IgG Titer Values (log) by Time and Outcome

|  | Antigen Targets |  |  |  |  |  |
| --- | --- | --- | --- | --- | --- | --- |
|  | S1 |  |  | S2 |  |  |
|  | Expired | Discharged | Difference | Expired | Discharged | Difference |
| Wk1 | 6.65 | 13.5 | **6.86\*** | 12.8 | 16.2 | **3.41\*** |
| Wk2 | 16.7 | 18.5 | 1.72 | 19.1 | 19.2 | 0.09 |
| Beyond Wk2 | 17.9 | 20.6 | 2.66 | 18.5 | 20.4 | 1.91 |
| Change from Wk1 to Wk2 | 10.1 | 4.95 | **-5.14\*** | 6.32 | 2.99 | **3.33\*** |
| Change from Wk1 to beyond Wk2 | 11.2 | 7.04 | **-4.20\*** | 5.73 | 4.23 | 1.50 |

\*Statistically Significant Differences in IgG Titer Values between Expired and Discharged groups at $\alpha=0.05$ .

While the above approaches helped us demonstrate significant differences in the early antibody response between patients who expired and those who were discharged, it did not tell us whether the antibody response trajectory itself can be predictive of the eventual outcome. To answer this question, we used a joint model (Model 2, Supplementary Materials) to relate the patient-specific antibody response to the eventual outcome. Specifically, we first fit a linear mixed model to the data to model the antibody response using time as a predictor and allowing for patient-specific deviations from the mean response at each time point. Then, we used the patient-specific predictions from this model as predictors in a probit regression model (Supplementary Materials) to predict the eventual outcome of death. The predicted patient-specific antibody levels were significant predictors of the outcome. The joint modeling approach produced models with AUC of 0.802 (95% CI: 0.647 -- 0.957) and 0.760 (95% CI: 0.578 -- 0.943) for spike protein domains S1 and S2 respectively (Figure 3), indicating that the patient-specific trajectories are highly predictive of the eventual outcome.

**Figure 3.**
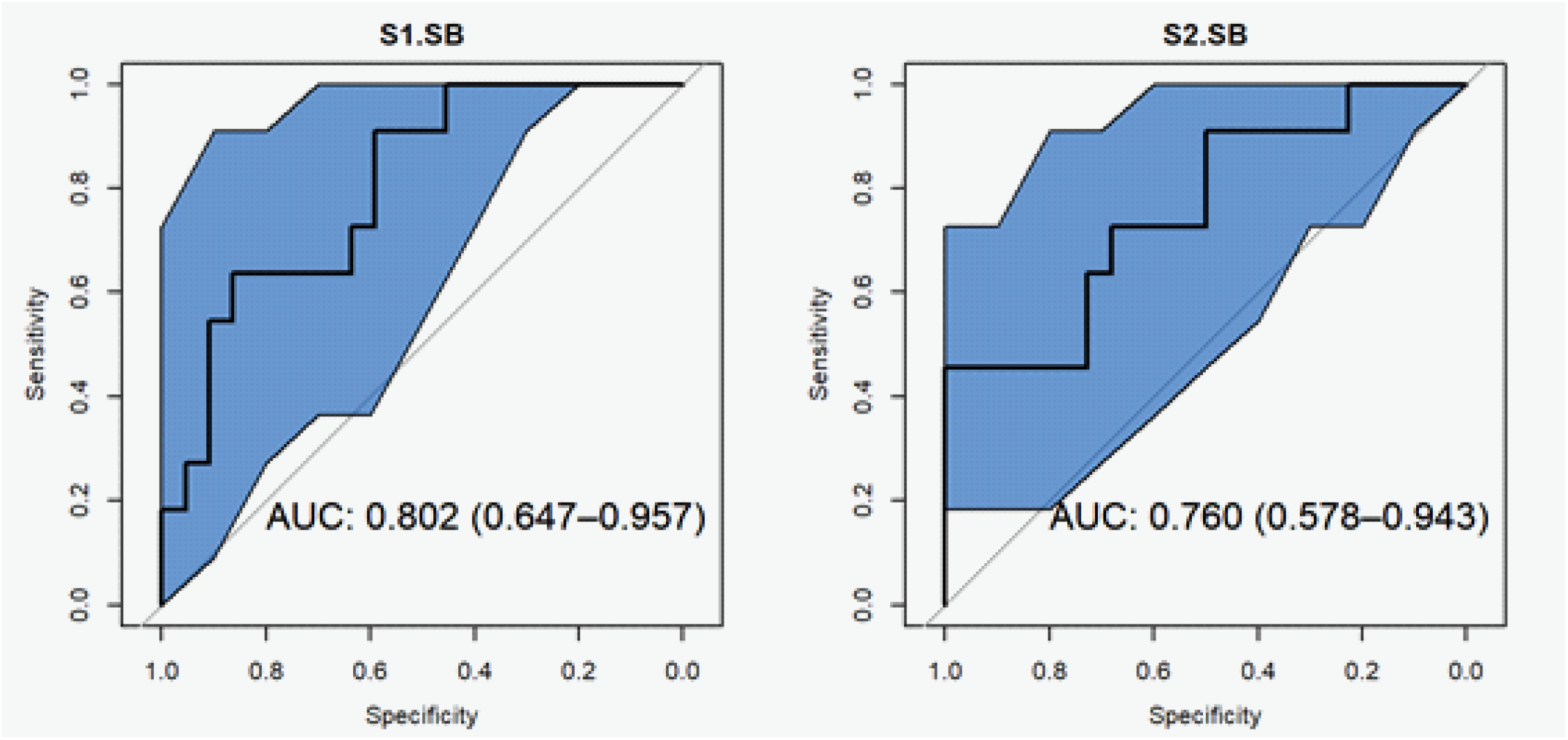
ROC Curves for Joint Model (Model 2) to Predict Death Using IgG Titer Values. Area under the curve (AUC) is presented within figure with 95% confidence intervals (shown in the blue regions) for specific IgG antibodies to S1 and S2, respectively.

Results for the linear mixed model and the joint model for other antibody isotypes, IgM and IgA, are less striking as compared to IgG and are available in the Supplementary Materials. There is also a sensitivity analysis excluding two patients in the Supplementary Materials.

## Discussion

We observed that lower levels of early antibody responses to several spike protein antigens correlated with poor outcomes. In addition, a joint model (Model 2) was developed, based on antibody trajectory, that was predictive of poor outcomes. This appears to differ with prior studies of SARS-CoV-1 (13) and SARS-CoV-2 (14) that noted a negative correlation with early antibody peak levels. These studies consider early seroconversion as <16 days or 8-14 days. When the actual timing of the results is compared, these studies are not in conflict with our data regarding early seroconversion at < 7 days from symptom onset. Although early antibody detection is seen in other studies (19) many assay formats have limited sensitivity (20) and may not detect early antibody, at low levels. Bead-based immunoassays can be highly sensitive (15, 21) allowing for early antibody detection. In addition, the bead-based assay described here had a similar binding pattern to other assays using ten panel samples in the First WHO SARS-COV-2 International Standard harmonization exercise (22).

This study has limitations, including the small number of patients and the evaluation of patients relatively early in the pandemic, with a high mortality. 10 out of 11 mortality outcomes were observed in males. Given our small sample size, we are not able to investigate if the relationship between antibody response and outcome was moderated by sex; However, there was a similar relationship when only male samples were evaluated (Supplementary Materials). Despite these limitations, a clear association between early antibody generation and survival is noted. These findings are consistent with other studies when matched for timeframe. These findings are robust across multiple Spike protein antigens and sensitivity analyses. This pattern was also observed with antibodies to RBD which are strongly correlated with neutralizing activity (23). A cut-point based maximum signal method had even stronger associations with early antibody levels (Supplementary Materials). Antibody data from past and future studies can be evaluated to further refine and confirm these findings; There may be other qualitative antibody characteristics associated with patient outcomes (24). Early antibody responses may relate to variability in time of infection to symptom onset and reflect other mechanisms related to disease onset.

Although the mechanisms for these findings need further elucidation, these results align well with clinical studies on therapeutic antibodies (2). These antibodies show benefit in outpatients with mild to moderate COVID-19, before further disease progression (3-5, 25, 26) whereas benefit of antibody treatment has not been observed in clinical trials in patients hospitalized due to COVID-19, and may be associated with worse clinical outcomes when administered to hospitalized patients with COVID-19 requiring high flow oxygen or mechanical ventilation (11, 12, 27). The importance of antibody early in disease also fits with data on early treatment using convalescent plasma (16, 17) but not later treatment (18) and also with the initial evaluations of vaccines that generate neutralizing antibodies. The exact stage of disease progression that may have clinical benefit from antibody therapy needs further study and may be dependent on the nature of the antibodies, additional interventions and other factors.

In developing an early antibody response as a predictor of outcomes, it is important to have sensitive method to reliably detect antibody early in the course of disease, such as the flow cytometry bead-based method we describe. Such an assay with appropriate modeling could be studied as an approach to predict the clinical course of patients and aid in selection of the appropriate therapeutic interventions.

## Materials and Methods

### Patient samples

Deidentified samples were obtained from Shady Grove Washington Adventist Hospital. Patients were hospitalized and diagnosed with COVID 19 by a PCR method. Onset of patient symptoms was in April through May 2020. Serial samples from 33 patients were evaluated; 11 of the patients expired and 22 were discharged. There were 4 to 16 consecutive collections per patient over a range of 0 to 42 days post-symptom onset. A variety of anti-coagulant tubes were used for blood collection including Heparin lithium, EDTA for plasma or serum. The patient manifestations, demographics and outcomes were blinded before evaluating sample antibody profiles detection and unblinded for correlation with outcomes after antibody levels were determined. Pre-COVID serum and plasma samples were retained from previous research projects of the lab. These were deidentified samples without additional information. COVID-19 patient samples were heat-treated at 60^°^C for one hour to de-activate the virus before performing the antibody assay.

This study was reviewed by the FDA Office of the Chief scientist and CBER Center Human Subject Protections (HSP) Liaison who determined that the study does not require FDA IRB review and approval because it is not research involving human subjects as defined in 45 CFR part 46 and it is not an FDA-regulated clinical investigation, as defined in 21 CFR part 56.

### Reagents

Functional CBA beads were purchased from BD Biosciences (San Jose, CA). Recombinant SARS COV2 antigens were purchased from commercial vendors: RBD-His was purchased from (Raybiotech, #230-30162) and (Sino Biological #40150-V08B2); S1 ECD-His (40591-V08H), S1D614G (40591-V08H3), S2 ECD-His (40590-V08B), S1+S2 ECD (40589-V08B1) were purchased from Sino Biological; Recombinant SARS-CoV-2 Nucleocapsid Protein (NP) was purchased from (Raybiotech).

Streptavidin-PE, streptavidin-BV421, anti-human IgG-PE, anti-human IgM-V450, and biotinylated anti-human IgA monoclonal antibodies were purchased from BD Biosciences (San Jose, CA). The specificity of these reagents was confirmed by ELISA and/or flow cytometry.

### Multiplexed SARS CoV2 Antigen Beads Array

Multiplexed SARS CoV2 recombinant antigen-coupled target beads and BSA control beads were prepared using sulfo-SMCC chemistry with Functional Bead Conjugation Buffer Set (BD Biosciences) according to manufacturer’s instructions. Conjugation of antigens and antibody binding were confirmed by flow cytometry using ELISA-confirmed rabbit anti-SARS CoV2 immune serum, SARS CoV1 immune rabbit serum, non-immunized control rabbit serum, as well as convalescent human COVID19 serum and pre-COVID19 human serum samples. Binding specificity was confirmed by free antigen inhibition of detection signal. In brief, serum samples were serially diluted with PBS buffer (containing 1% BSA and 2mM EDTA) from 1:10 up to 1:100,000,000, mixed with SARS-CoV-2 bead arrays, with and without free SARS-CoV-2 antigens, incubated at 4°C with shaking overnight; washed with PBS buffer, developed with anti-human Ig isotype antibodies, and detected by flow cytometry (BD LSRFortessa™). Flow data files were analyzed using FlowJo software (FlowJo, LLC, Ashland, OR).

The multiplexed beads array method was demonstrated to sensitively and specifically detect antibody signals in rabbit anti-SARS-CoV-2 immune serum, convalescent human COVID-19 serum, and serum samples from patients with PCR-confirmed COVID19.

The assay was also used to participate in the Establishment of the 1st WHO International Standard and Reference Panel for anti-SARS-CoV-2 antibody (22). Using our flow-cytometry based assays for the detection of IgA, IgG and IgM against SARS-CoV-2 antigens (E, M, N, RBD, S1, S2 and S1+S2ECD) and expressing the IgG data as relative to the candidate International Standard IgG with an assigned arbitrary unitage of 1000 IU/mL, the binding pattern obtained with the ten panel samples, was similar to the pattern obtained with all of the 27 neutralization assays that participated in the harmonization exercise (Figure 2 and Figure 6 in the WHO online report).

### Detection of anti-CoV2 antibody

Detection for anti-CoV2 antibodies in heat-inactivated serum or plasma samples was done using 96 well U-shaped plates for high-throughput runs. Serum/plasma samples were serially diluted 1:100, 1:1,000, 1;10,000 and 1:100,000 for detection of IgG and IgA, and 1:20, 1:80, 1:320, 1:1,280 for detection of IgM. For every 96-well plate, ∼ 2.4 x 10^6^ beads containing targets and control beads were added to 2 mL serum enhancement buffer (supplied in the BD human CBA kits). Target beads and control beads were mixed in total of 9 mL buffer (2 mL serum treatment buffer, 2 mL beads capture buffer, 5 mL sample diluent). Then 80µL beads and 80 µL of serum sample were added per well (96-U plate), leading to another 2-fold sample dilution, incubated overnight at 4^°^C with shaking and washed twice. Ten µL/well of anti-human IgGFc-PE, 1 µL/well of anti-human IgA-bio (and further developed by streptavidin-BV421) or 2 µL/well anti-human-IgM-V450 were added and after 1 hour, the wells were washed twice by centrifugation and analyzed by flow cytometry HTS run. Final sample dilutions were 1:200, 1:2,000, 1;20,000 and 1:200,000 for detection of IgG and IgA, and 1:40, 1:160, 1:640, 1:2,560 for detection of IgM. Fourteen pre-COVID serum samples were used as negative controls. Flow cytometry data were analyzed with FlowJo software (FlowJo, LLC); More than 10,000 signal events were collected per sample. Single bead populations were gated by FSC-SSC. Target beads and control beads were separated by APC & ACP-Cy7 fluorescence intensity and properly compensated for effective display. PE & V450 or BV421 fluorescence intensity of target beads and control beads were exported to Excel files and further analyzed for antibody levels.

Antigen-specific antibody levels (i.e., extrapolated titer value) are the maximum inverse dilution factor where the MFI of titration curves plateau. Due to limited titrations, antigen-specific antibody levels (i.e., extrapolated titer values) were calculated based on multiplying the inverse of each sample dilution by an additional inverse dilution factor and using the maximum value. The additional inverse dilution factor was calculated based on a linear regression (Excel Forecast function) between MFI and dilution. The regression used the linear segment of a selected sample titration curve vs. MFI, on each plate, as a standard curve. An alternative set of antigen-specific antibody levels were calculated based on the maximal signal of inverse dilution factor x median fluorescent intensity (MFI) for each titration; in this calculation, MFI must meet a threshold-criteria based on the average + 3SD of 14 pre-COVID 19 samples. All the calculated antibody level values were log transformed for further analysis.

### Antibody Level Analysis and Predictive Modeling for Outcome

Log2 transformed antibody levels over time, starting from the earliest sample collection date after onset, in the expired and discharged patients were plotted. The trajectories of antibody levels are displayed for hospitalized patients who expired and who were discharged alive. For patients who expired, the earliest sample was at Day 0 and the latest sample was at Day 30, with a median of 6 samples per patient. For patients who were discharged, the earliest sample was at day 0 and the latest sample was at Day 42, with a median of 6 samples per patient.

As there were limited sequential sample days for any given patient, we tested for differences across expired and discharged groups using antibody levels at week one, week two and after week two. For patients with more than one sample within a week, we used the median of the available antibody levels. Density and box plots were used to display the distribution of antibody levels at week one, two and after week two, along with results from a t-test comparing the means of the distributions for different proteins.

We also fit a linear mixed model (Model 1), accounting for patient-level correlation, to the data to model the antibody response using time, expired / discharged status and the interaction between time and expired / discharged status as predictors. Specifically, the mean antibody levels at week 1, week 2 and after week 2 were estimated for the expired group and the discharged group. The estimated antibody titer value differences between expired group and the discharged group were calculated and underwent statistical test. The mean increase antibody titer of week 2 and after week 2 from week one value was also calculated for each patient group to compare antibody response.

To determine whether the antibody response trajectory itself can be predictive of the eventual outcome, we used a joint model (Model 2) to relate the patient-specific antibody response to the eventual outcome. Specifically, we first fit a linear mixed model to the data to model the antibody levels using time as a predictor and allowing for patient-specific deviations from the mean response at each time point. Then, we used the patient-specific predictions from this model as predictors in a probit regression model to predict the eventual outcome of Death (or expired status). The predicted patient-specific antibody levels were used as predictors of the outcome as in model 1 and the joint modeling approach Receiver Operating Characteristic (ROC) curves were plotted and under the curve (AUC) calculated within figure with 95% confidence intervals. Details on the modeling are in the Supplementary Materials under Statistical Modeling.

## Data Availability

Contact the corresponding authors for data and materials; Materials are commercially available except for patient samples; Patient samples may have limited availability.

## Acknowledgements

We would like to thank Daniela Verthelyi, Mohan Manangeeswaran, Amy Rosenberg, Adam Fisher, Adam Sherwat, John Farley and Sau Lee for critical review of the manuscript. We also thank Hana Golding, Emily Braunstein and Carolyn Wilson for coordinating of clinical samples. We would like to thank Mat Soukup and Yong Ma for critical review of statistical analysis. We thank JuMe Park, Sujata Bupp and Hanxia Huang for lab and technical support.

For providing the patient samples we thank: James Rost, CMO, White Oak Medical Center, and Norton Elson, AHC Medical Director Quality and Clinical Effectiveness of Washington Adventist Medical HealthCare; Nicolas Cacciabeve, Laboratory Medical Director of Advanced Pathology Associates; Rob San Luis, Hollie Genser, Demetra Collier, Meaza Belay, Genevieve Caoili, Zanetta E. Morrow and Bryana Streets of Quest Diagnostics.

Steven Kozlowski and Zhao-Hua Zhou are members of the Office of Pharmaceutical Quality Center of Excellence for Infectious Diseases and Inflammation at CDER, FDA.

This article reflects the views of the authors and should not be construed to represent FDA’s views or policies.

## Funding

This study was funded with FDA MCM resources.

## Author Contributions

Conception of the work: ZZ, SK

Performing experiments: ZZ

Data analysis and interpretation: ZZ, SD, ML, SLK, SK

Statistical analysis: SD

Drafting and revision of the manuscript: ZZ, SD, ML, SLK, SK

## Competing interests

The authors declare no conflicts of interest.

## Supplementary Materials

Statistical Modeling

Assay Characteristics Figure S1, S2, S3

IgG Levels at Week 2 and After Week 2 Post-onset Comparisons Figure S4, S5, Table S1

IgG Analysis Using IgG Cut-point MFI Method Figure S6

IgG Cut-point MFI Week 1 Post-onset Comparison Figure S7

IgG Cut-point MFI Modeling Table S2, Figure S8

IgM Analysis

Anti-SARS CoV2 IgM Antibody Kinetics Figure S9

IgM Week 1 Post-onset Comparison Figure S10 IgM Modeling Table S3, Figure S11

IgA Analysis

Anti-SARS CoV2 IgA Antibody Kinetics Figure S12

IgA Week 1 Post-onset Comparison Figure S13

IgA Modeling Table S4, Figure S14

IgG Sensitivity Analysis (exclude 1000 and 1007)

Anti-SARS CoV2 IgG Sensitivity Analysis Kinetics Figure S15

IgG Sensitivity Analysis MFI Week 1 Comparison Figure S16

IgG Sensitivity Analysis Modeling Table S5, Figure S17

IgG Sensitivity Analysis (Males)

Anti-SARS CoV2 IgG Sensitivity Analysis Kinetics Figure S18

IgG Sensitivity Analysis MFI Week 1 Comparison Figure S19

IgG Sensitivity Analysis Modeling Table S6, Figure S20

## Notes

### Competing Interest Statement

The authors have declared no competing interest.

