## Supplementary Materials for "Early Antibody Responses Associated with Survival in COVID19 Patients"

### Contents


**Statistical Modeling** ... 2

**Model 1** ... 2

**Model 2** ... 3

**Assay Characteristics** ... 4

**IgG Levels at Week 2 and After Week 2 Post-onset Comparisons** ... 7

**IgG Analysis Using IgG Cut-point MFI Method** ... 9

**IgG Cut-point MFI Week 1 Post-onset Comparison** ... 10

**IgG Cut-point MFI Modeling** ... 10

**IgM Analysis** ... 12

**Anti-SARS CoV2 IgM Antibody Kinetics** ... 12

**IgM Week 1 Post-onset Comparison** ... 13

**IgM Modeling** ... 13

**IgA Analysis** ... 15

**Anti-SARS CoV2 IgA Antibody Kinetics** ... 15

**IgA Week 1 Post-onset Comparison** ... 16

**IgA Modeling** ... 16

**IgG Sensitivity Analysis (exclude 1000 and 1007)** ... 18

**Anti-SARS CoV2 IgG Sensitivity Analysis Kinetics** ... 18

**IgG Sensitivity Analysis MFI Week 1 Comparison** ... 19

**IgG Sensitivity Analysis Modeling** ... 19

**IgG Sensitivity Analysis (Males)** ... 21

**Anti-SARS CoV2 IgG Sensitivity Analysis Kinetics** ... 21

**IgG Sensitivity Analysis MFI Week 1 Comparison** ... 22

**IgG Sensitivity Analysis Modeling** ... 22

### Statistical Modeling

#### Model 1

To examine the relationship between antibody values and outcomes we fit the following linear mixed model for Antibody Response at time $j$ for patient $i$ with outcome $Outcome_i$:

$$
\begin{aligned}
Antibody\ Response_{ij} \\
&= \beta_0 + \beta_1 I(j = Week_2) + \beta_2 I(j = Beyond\ Week_2) \\
&+ I(Outcome_i = Expired) \\
&* [\beta_3 + \beta_4 I(j = Week_2) + \beta_5 I(j = Beyond\ Week_2)] + b_i + \epsilon_{ij}
\end{aligned}
$$

In the above model, $\beta_0$ and $\beta_3$ represent the mean response (i.e., antibody titer value) at Week 1 for discharged and expired patients, respectively. The mean increase from Week 1 to Week 2, and Week 1 to after Week 2, is captured by $\beta_1$ and $\beta_2$, respectively, in discharged patients and by $\beta_4$ and $\beta_5$, respectively, in expired patients. We account for correlation between observations at different time points for each patient by including a patient-specific random effect $b_i$. The random effects for patients are assumed to arise from a normal distribution, i.e., $b_i \sim N(0, \tau)$.

### Model 2

To examine the predictive ability of patient-specific antibody response trajectory, we used a joint modeling approach[1]. We first model the antibody response using a linear mixed model, similar to model 1 but without including outcome as a predictor, as follows:

$$Antibody\ Response_{ij} = \beta_0 + \beta_1 I(j = Week_2) + \beta_3 I(j = Beyond\ Week_2) + b_{i0} + b_{i1} I(j = Week_2) + b_{i2} I(j = Beyond\ Week_2) + \epsilon_{ij}$$

Here, random effects $b_{i0}, b_{i1}, b_{i2}$ represent patient-specific deviations from the population mean of response at Week 1, increase from Week 1 to Week 2, and Week 1 to after Week 2, respectively. The estimates of the patient-specific deviations, $\hat{b}_{i0}, \hat{b}_{i1}, \hat{b}_{i2}$, obtained from fitting the above model are then used as predictors in a probit regression model to predict the eventual outcome as follows:

$$Prob(Outcome_i = Expired) = \phi[\gamma_0 + \gamma_1 \hat{b}_{i0} + \gamma_2 \hat{b}_{i1} + \gamma_3 \hat{b}_{i2}]$$

Here, $\phi$ denotes the cumulative distribution function of the standard normal distribution. The predictive ability of the model is measured using the area under the receiver operating characteristic curve (AUC).

[1] Albert PS. A linear mixed model for predicting a binary event from longitudinal data under random effects misspecification. Statist Med. 2012;31(2):143-154.

### Assay Characteristics

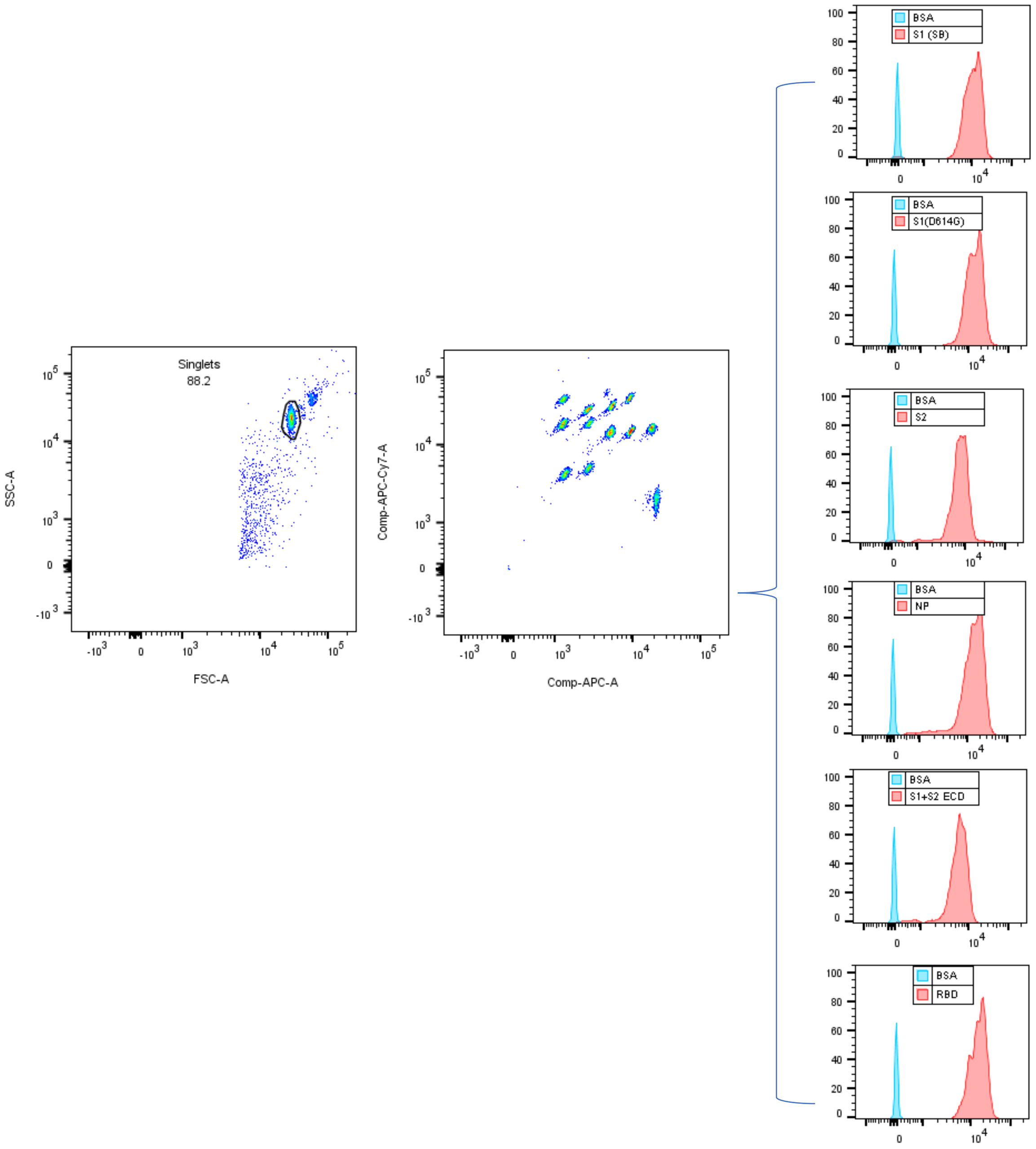


**Figure S1 Multiplexed beads array for COV2 antibody and flow cytometry analysis.** The singlet beads population were gated based on FSC-SSC display, followed by compensated APC-APC-Cy7 display to show different SARS-CoV2 antigen component-coated target and BSA-control beads. Histograms display overlays of antibody binding signals to antigen-target beads vs BSA beads as indicated.

### Assay Sensitivity and Specificity

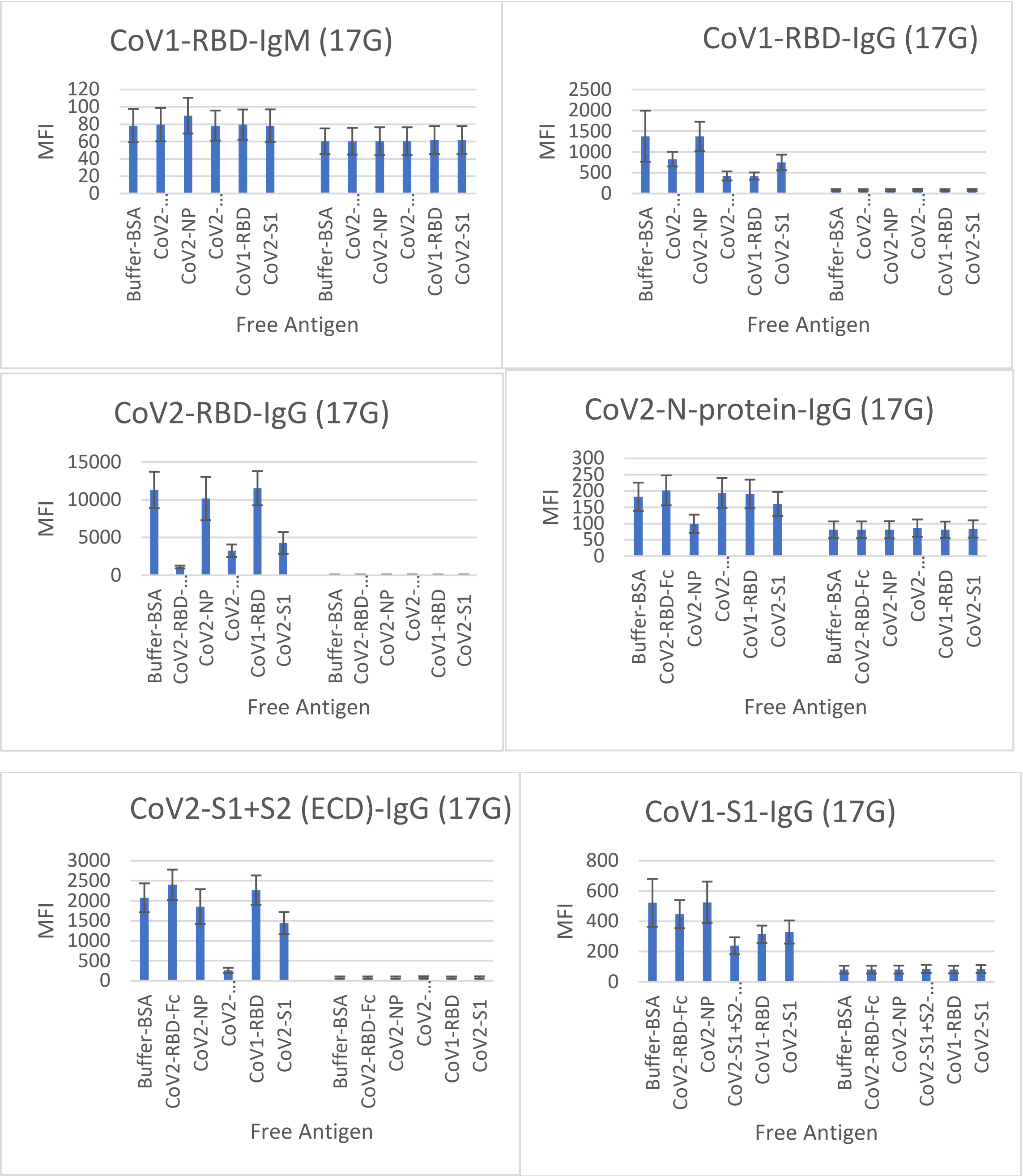


**Figure S2 Assay Sensitivity and Specificity.** Serum from a COVID19 patient (17G, left side of panels) and a pre-COVID control sample (right side of panels) were incubated with an array of SARS-CoV1 and CoV-2 antigen coated beads, in the presence of free antigens or BSA as indicated. Specific binding signals (MFI) were compared indicating 17G but not the pre-COVID sample contained specific IgG antibodies to COV2-RBD, COV2-N, COV2-S1+S2 (ECD), and cross-reactive to CoV1-RBD and CoV1-S1 (with much less signal intensity as compared to CoV2 antigens). The signals could be inhibited by free antigens as coated on the target beads or free antigens with overlapping epitopes as coated on the target beads.

### Antibody Titer calculation with on-plate standard curve

### Figure S3 Example of MFI vs dilution factor chart for antibody to RBD and BSA

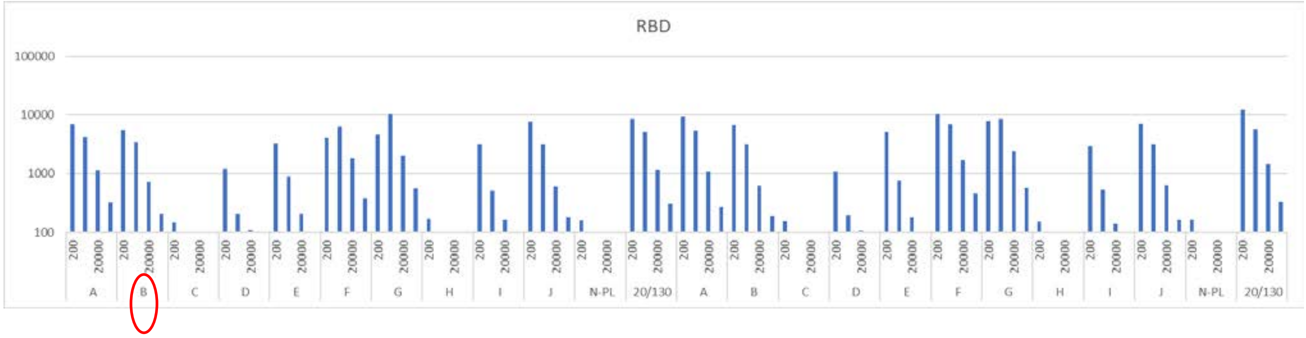


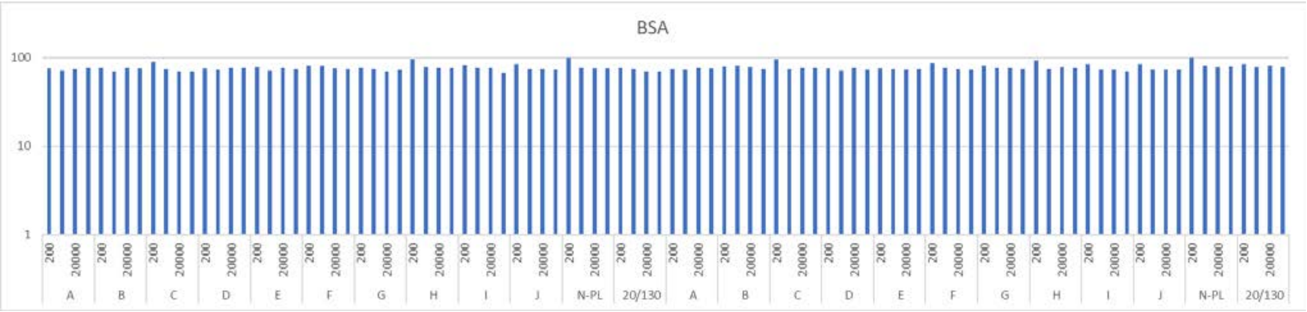


Sample B was chosen as on-plate assay standard to calculate and normalize titer values of all the samples on the same plate (Threshold was at MFI=205)

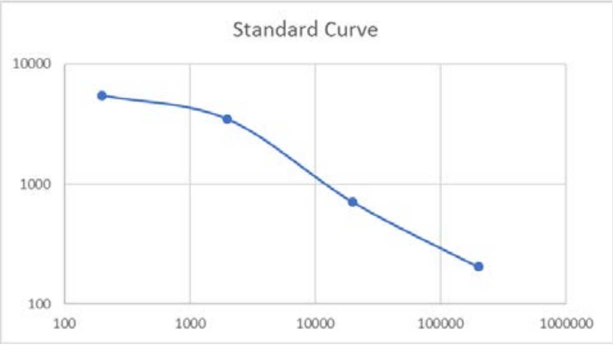


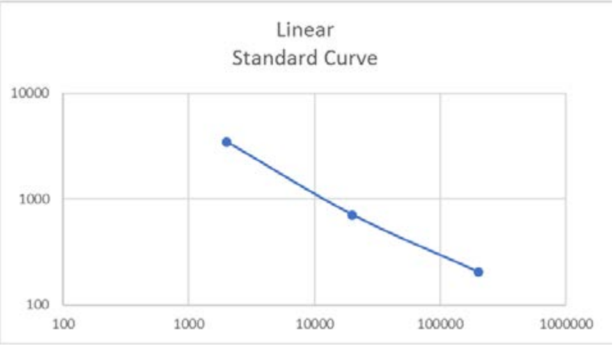


Calculated anti-RBD antibody titer value based on linear regression extrapolation

| Sample | A | B | C | D | E | F | G | H | I | J | N-PL | 20/130 |
|---|---|---|---|---|---|---|---|---|---|---|---|---|
| Titer | 215675 | 133990 | 0 | 5759 | 25507 | 349687 | 709409 | 0 | 17137 | 119137 | 0 | 244130 |

### IgG Levels at Week 2 and After Week 2 Post-onset Comparisons

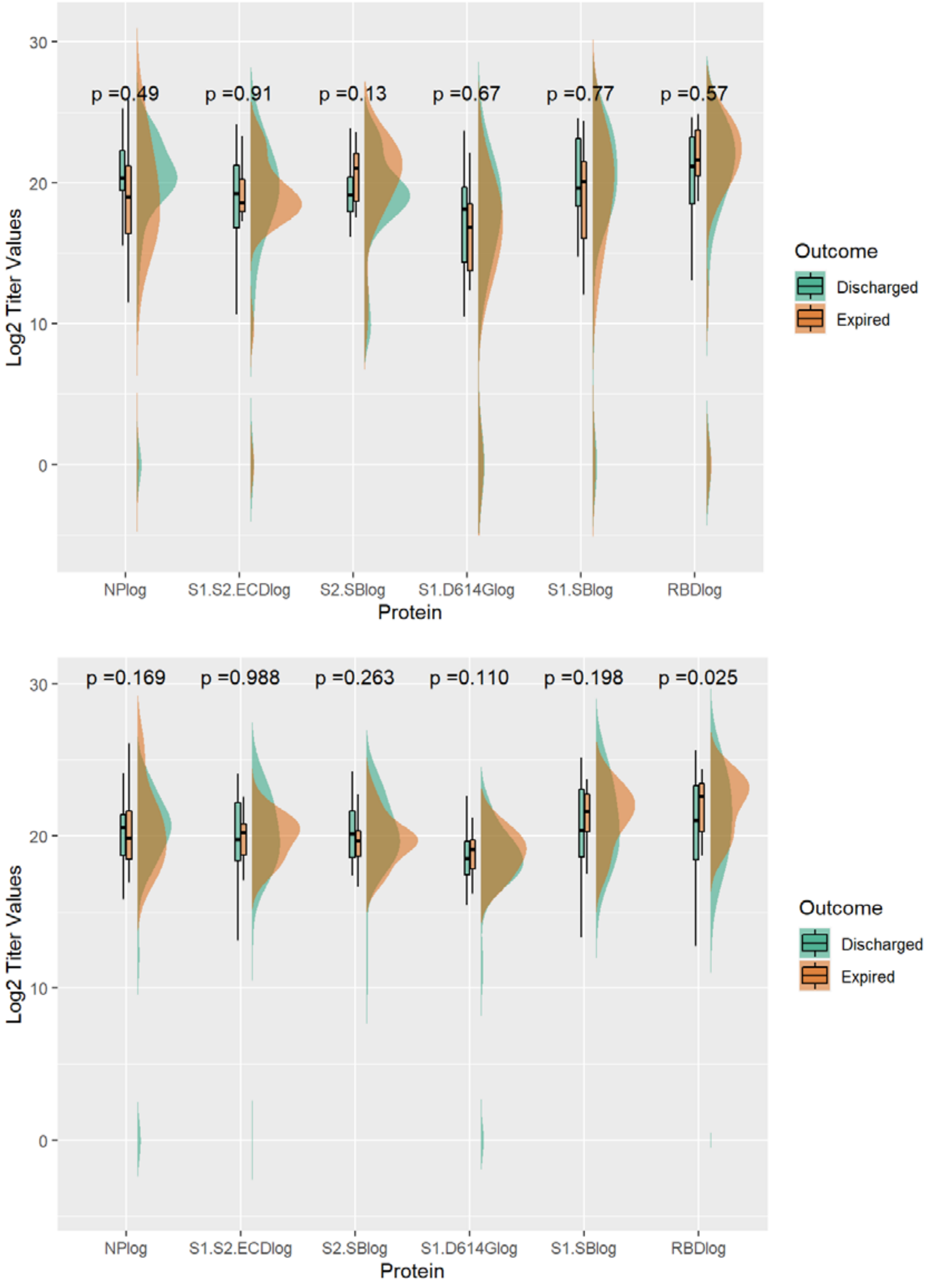


**Figure S4** Distribution of IgG antibody levels (log transformed) at week 2 (top) and after week 2 (bottom) post onset in expired and discharged groups with t-test comparison p-value.

**Table S1. Estimates from Linear Mixed Model (Model 1) of Difference between Discharged and Expired Groups in other IgG Antibody Levels (log) by Time**

| | Average Difference between Discharged and Expired groups | | | |
|---|---|---|---|---|
| | NP | S1S2ECD | S1D614 | RBD |
| Week1 | 2.71 | 2.62 | 4.89* | 4.61 |
| Week2 | 2.07 | 0.18 | 1.73 | 1.01 |
| Beyond Week2 | 1.79 | 1.78 | 1.44 | 2.32 |
| Change from week1 to week2 | -0.64 | -2.44* | -3.16 | -3.60 |
| Change from week1 to beyond week2 | -0.92 | -0.84 | -3.46* | -2.29 |
| *Statistically Significant Differences in IgG Titer Values between Discharged and Expired groups at $\alpha = 0.05$ | | | | |

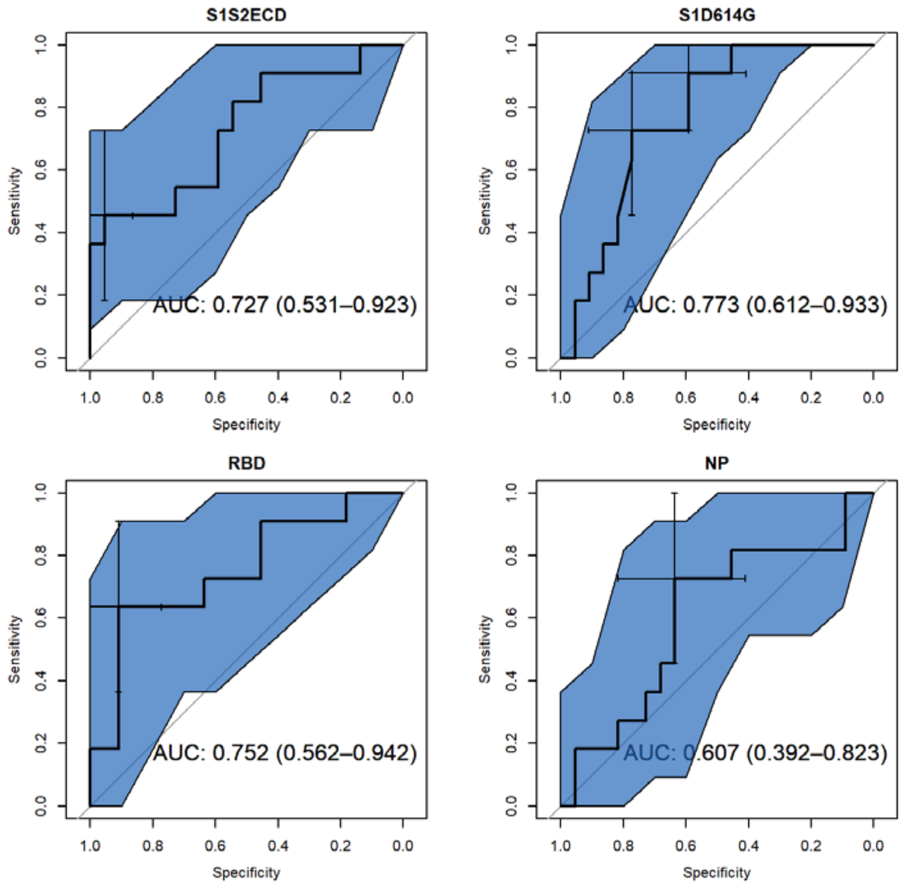


**Figure S5 Cut-point MFI.** ROC curves for joint model (model 2) to predict death using IgG antibody levels; area under the curve (AUC) is presented within figure with 95% confidence intervals.

### IgG Analysis Using IgG Cut-point MFI Method
### Anti-SARS CoV2 IgG Cut-point MFI Antibody Kinetics

#### Analysis with Antibody level expressed by maximum signal of MFI x dilution factor for MFI > 3SD + Pre-COVID baseline

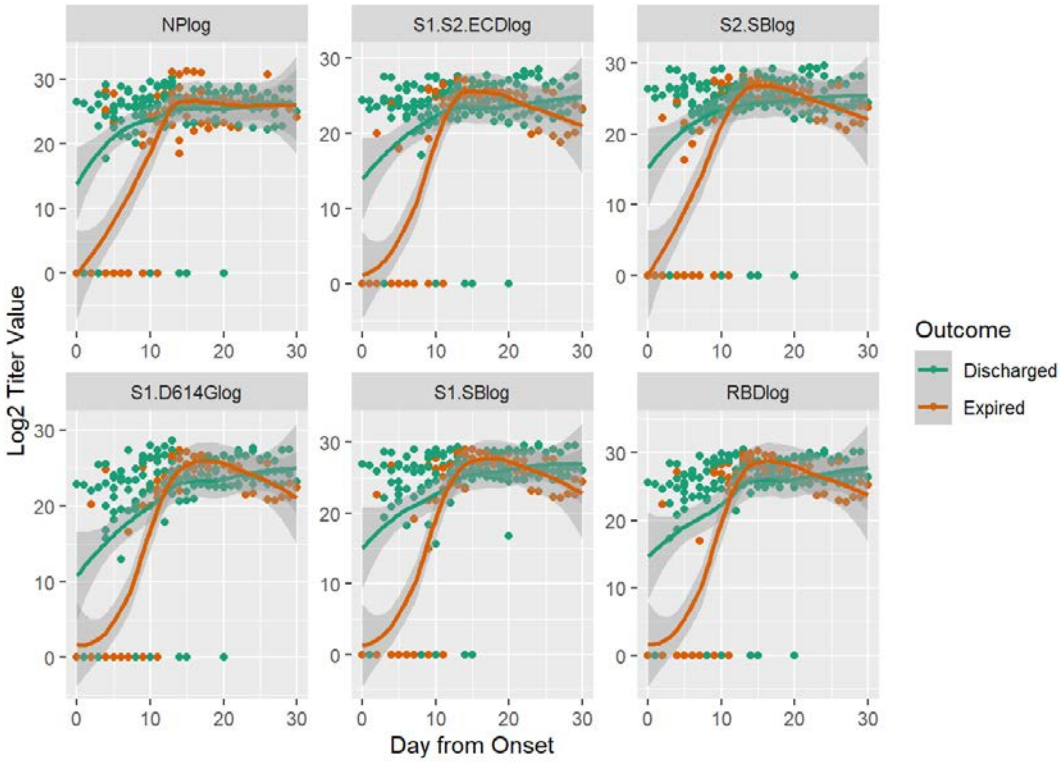


**Figure S6 Anti-SARS CoV2 IgG Cut-point MFI Antibody Kinetics.** Observed IgG antibody titer values (log transformed) in expired and discharged groups of patients by protein antigen. Points are observed values for each patient at corresponding day from onset; lines are smoothed regression lines fit to the observed data with 95% confidence interval bands.

### IgG Cut-point MFI Week 1 Post-onset Comparison

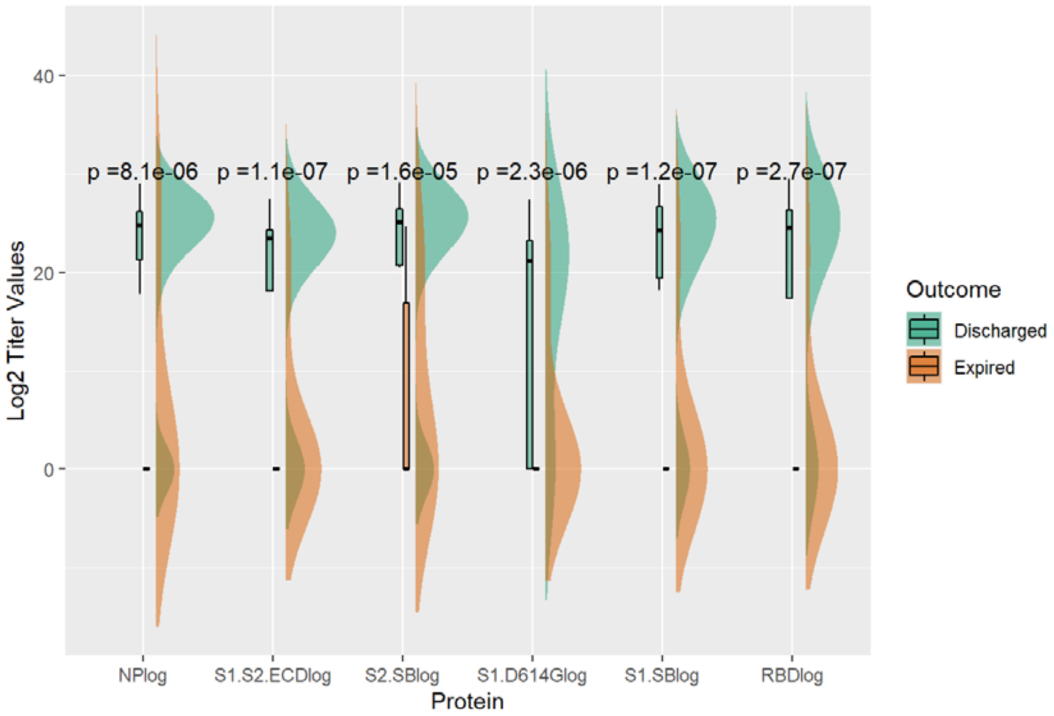


**Figure S7. Comparison of Week1 (0 to 7 days) After Onset Anti-SARS COV2 IgG Antibody Titers Between Discharged and Expired Groups using IgG Cut-point MFI.** Antibody Distribution of IgG titer values (log transformed) at week 1 post onset in expired and discharged groups with t-test comparison p-value.

### IgG Cut-point MFI Modeling

**Table S2. Estimates from Linear Mixed Model (Model 1) of Difference between Discharged and Expired Groups in IgG Antibody Levels (log) by Time**

| | Average Difference between Discharged and Expired groups | | | | | |
|---|---|---|---|---|---|---|
| | NP | S1 | S2 | S1S2ECD | S1D614 | RBD |
| Week1 | 9.59* | 11.1* | 8.40* | 10.0* | 7.59* | 9.38* |
| Week2 | 3.75 | 1.98 | 2.90 | 2.69 | 2.08 | 1.83 |
| Beyond Week2 | 1.99 | 3.18 | 2.62 | 1.99 | 2.26 | 2.17 |
| Change from week1 to week2 | -5.84* | -9.12* | -5.50* | -7.30* | -5.51* | -7.55* |
| Change from week1 to beyond week2 | -7.61* | -7.91* | -5.78* | -8.01* | -5.33* | -7.21* |
| *Statistically Significant Differences in IgG Titer Values between Discharged and Expired groups at $\alpha = 0.05$ | | | | | | |

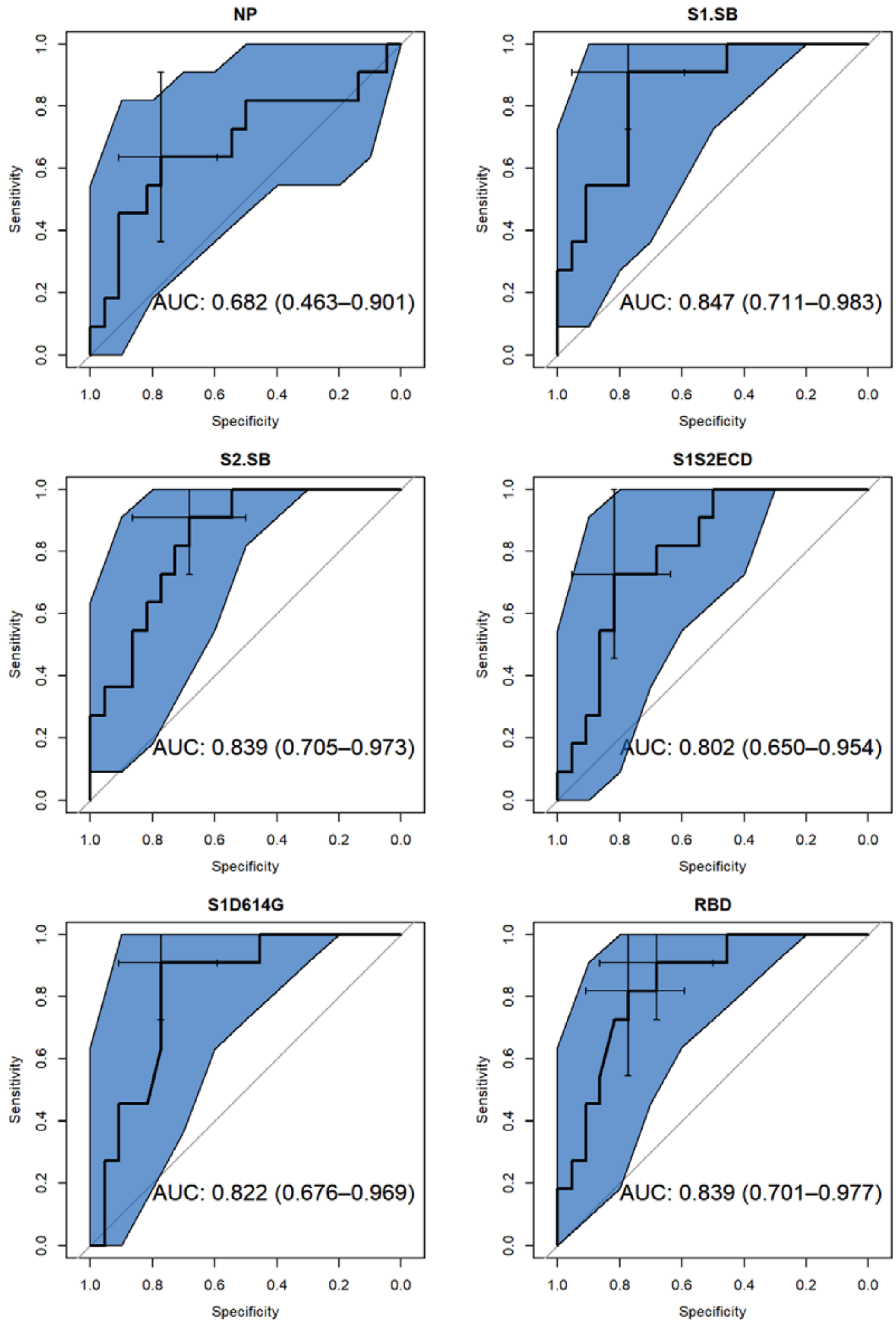


**Figure S8 Cut-point MFI.** ROC curves for joint model (model 2) to predict death using IgG antibody levels; area under the curve (AUC) is presented within figure with 95% confidence intervals.

### IgM Analysis

#### Anti-SARS CoV2 IgM Antibody Kinetics

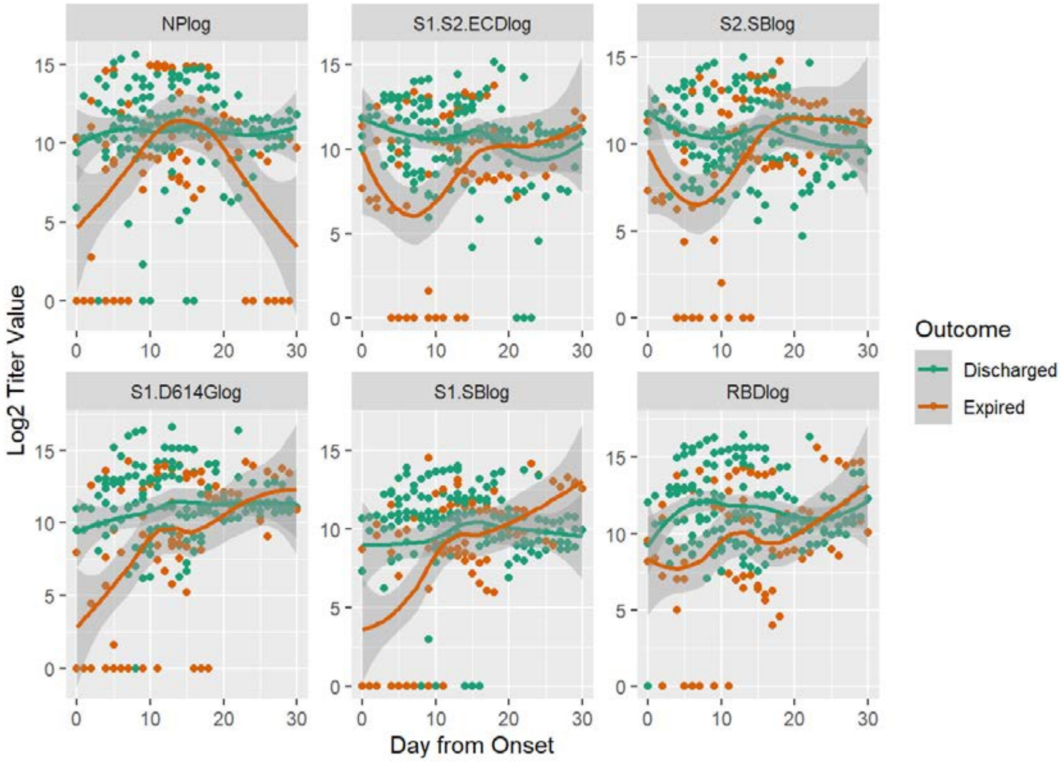


**Figure S9.** Observed IgM antibody titer values (log transformed) in expired and discharged groups of patients by protein antigen. Points are observed values for each patient at corresponding day from onset; lines are smoothed regression lines fit to the observed data with 95% confidence intervals.

### IgM Week 1 Post-onset Comparison

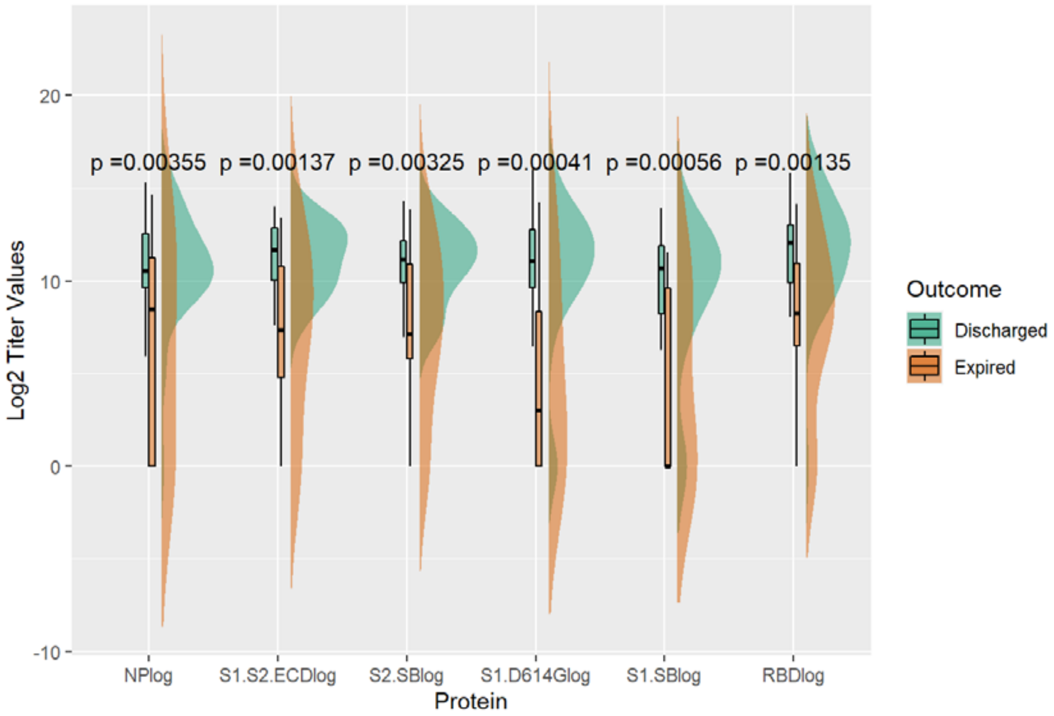


**Figure S10.** Distribution of IgM antibody levels (log transformed) at week 1 post onset in expired and discharged groups with t-test comparison p-value.

### IgM Modeling

#### Table S3. Estimates from Linear Mixed Model (Model 1) of Difference between Discharged and Expired Groups in IgM Antibody Levels (log) by Time

| | Average Difference between Discharged and Expired groups | | | | | |
|---|---|---|---|---|---|---|
| | NP | S1 | S2 | S1S2ECD | S1D614 | RBD |
| Week1 | 2.05 | 2.54 | 3.08* | 3.91* | 4.13* | 3.14* |
| Week2 | 0.70 | 0.42 | 1.27 | 1.52 | 1.42 | 1.33 |
| Beyond Week2 | 2.57 | 1.73 | 1.86 | 2.00 | 3.06* | 3.40* |
| Change from week1 to week2 | -1.35 | -2.12* | -1.81* | -2.38* | -2.71* | -1.81* |
| Change from week1 to beyond week2 | 0.52 | -0.81 | -1.22* | -1.91* | -1.07 | 0.27 |
| *Statistically Significant Differences in IgM Titer Values between Expired and Discharged groups at $\alpha = 0.05$ | | | | | | |

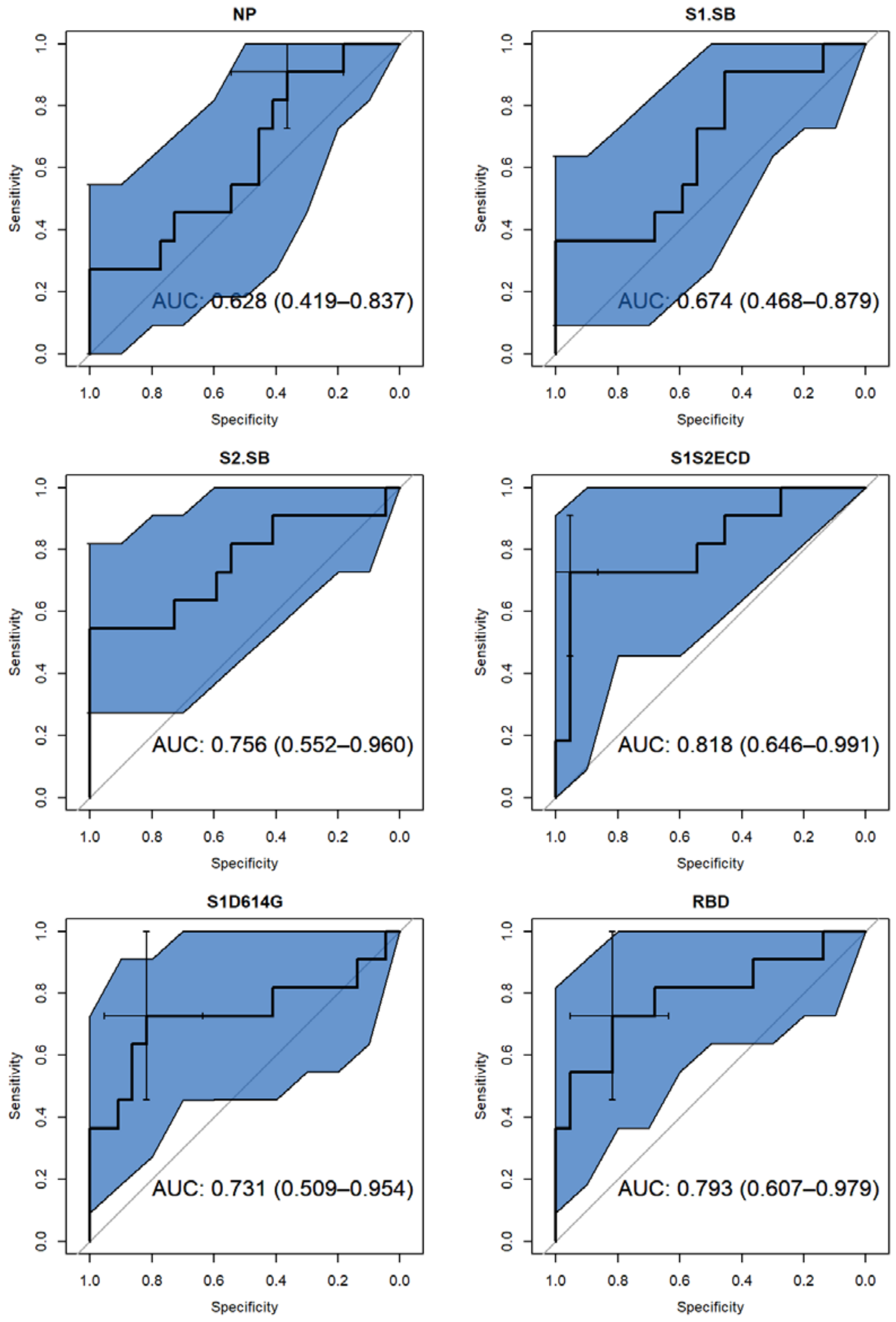


**Figure S11.** ROC curves for joint model (model 2) to predict death using IgM antibody levels; area under the curve (AUC) is presented within figure with 95% confidence intervals.

### IgA Analysis

#### Anti-SARS CoV2 IgA Antibody Kinetics

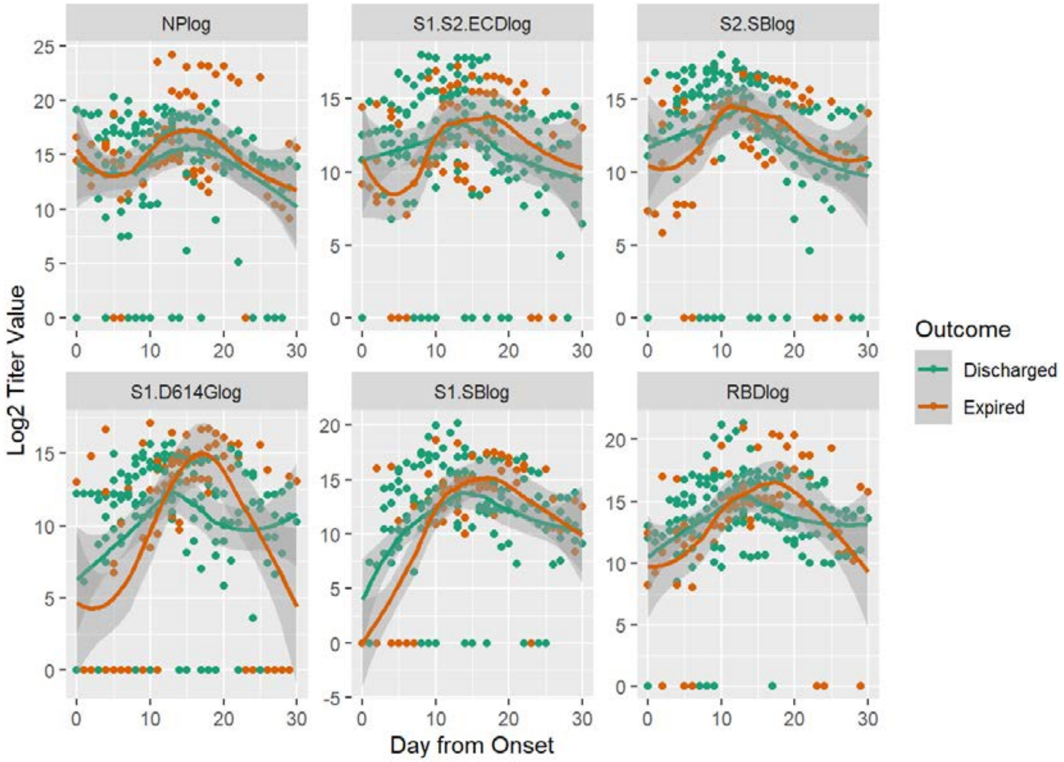


**Figure S12.** Observed IgA antibody levels (log transformed) in expired and discharged groups of patients by protein antigen. Points are observed values for each patient at corresponding day from onset; lines are smoothed regression lines fit to the observed data with 95% confidence intervals.

### IgA Week 1 Post-onset Comparison

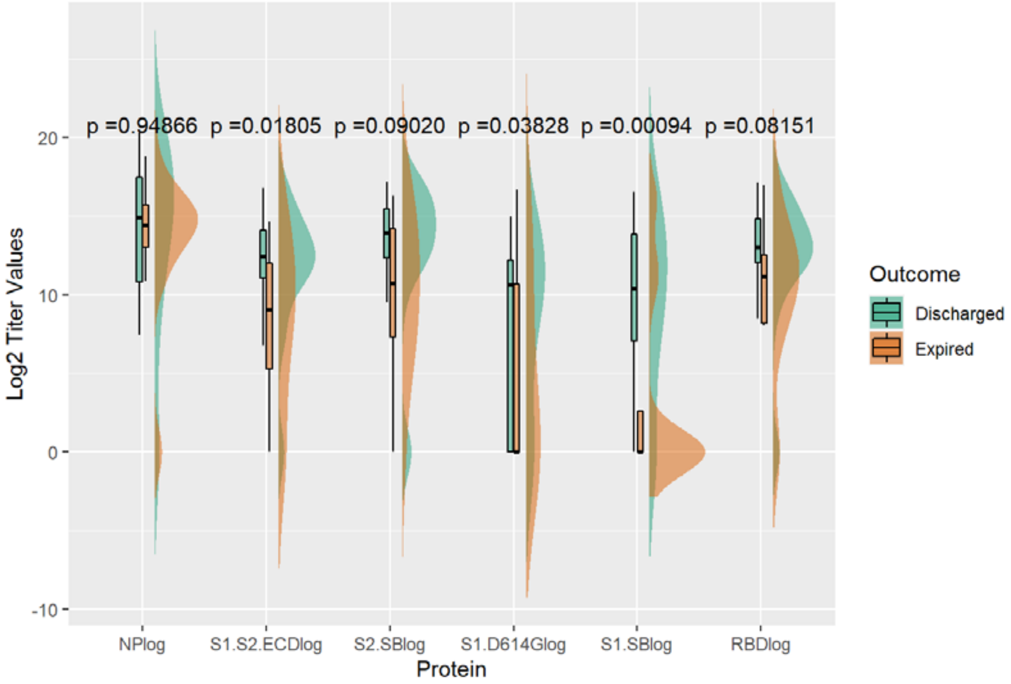


**Figure S13.** Distribution of IgA antibody levels (log transformed) at week 1 post onset in expired and discharged groups with t-test comparison p-value.

### IgA Modeling

#### Table S4. Estimates from Linear Mixed Model (Model 1) of Difference between Discharged and Expired Groups in IgA Antibody Levels (log) by Time

| | Average Difference between Discharged and Expired groups | | | | | |
|---|---|---|---|---|---|---|
| | NP | S1 | S2 | S1S2ECD | S1D614 | RBD |
| Week1 | -0.57 | 4.42* | 1.17 | 2.83 | 1.11 | 1.43 |
| Week2 | -2.02 | -1.35 | -1.20 | -1.31 | 0.25 | -0.43 |
| Beyond Week2 | -0.63 | -0.04 | 0.02 | -0.92 | -0.09 | -0.04 |
| Change from week1 to week2 | -1.45 | -5.77* | -2.37 | -4.15* | -0.86 | -1.86 |
| Change from week1 to beyond week2 | -0.06 | -4.46* | -1.15 | -3.75* | -1.19 | -1.48 |
| *Statistically Significant Differences in IgA Titer Values between Expired and Discharged groups at $\alpha = 0.05$ | | | | | | |

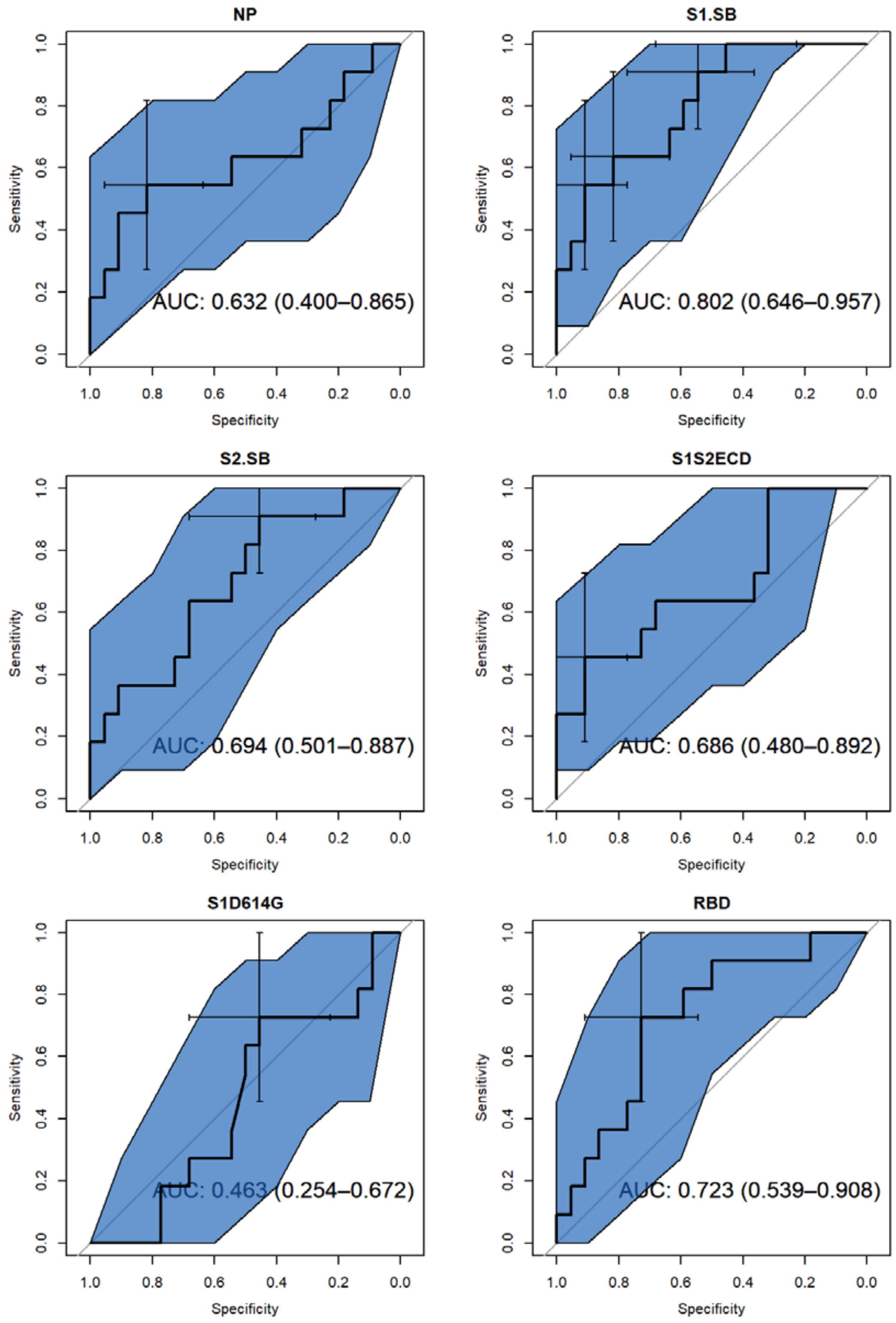


**Figure S14.** ROC curves for joint model (model 2) to predict death using IgM antibody levels; area under the curve (AUC) is presented within figure with 95% confidence intervals.

### IgG Sensitivity Analysis (exclude 1000 and 1007)

For two patients there was a potential misclassification of some samples based on a discrepancy between logs. To rule out any potential impact, analyses were run completely removing both of those patients. Exclusion of those patients had no impact on the results.

### Anti-SARS CoV2 IgG Sensitivity Analysis Kinetics

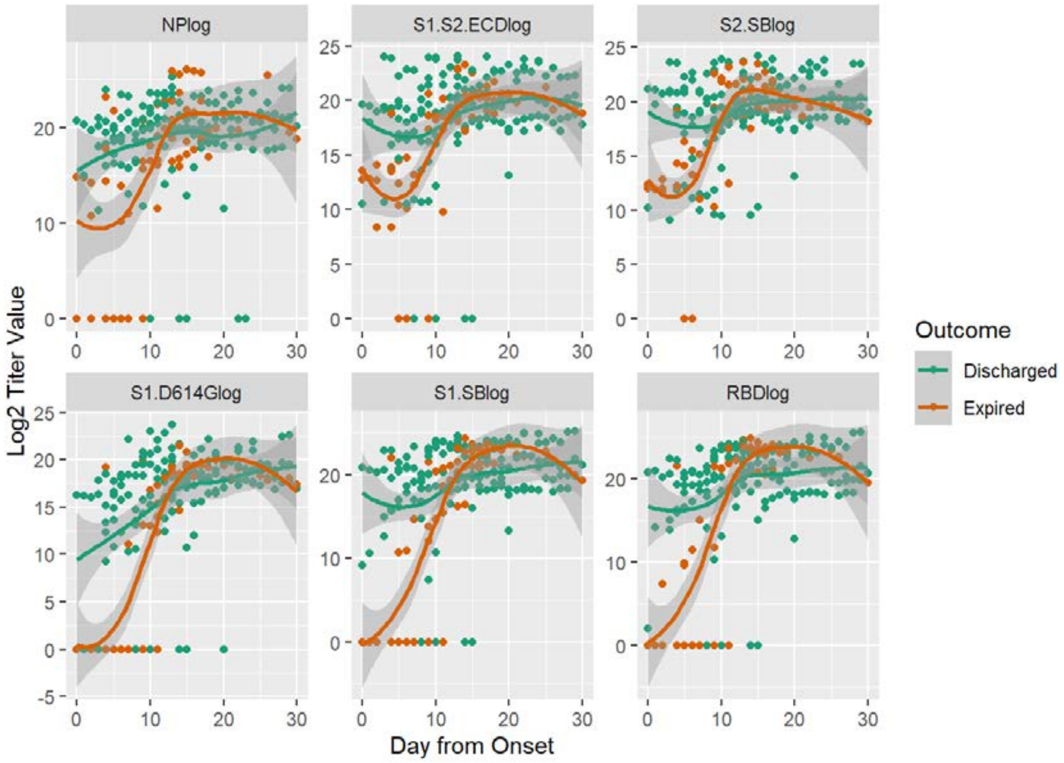


**Figure S15 Anti-SARS CoV2 IgG Kinetics (exclude 1000 and 1007).** Observed IgG antibody titer values (log transformed) in expired and discharged groups of patients by protein antigen. Points are observed values for each patient at corresponding day from onset; lines are smoothed regression lines fit to the observed data with 95% confidence intervals.

### IgG Sensitivity Analysis MFI Week 1 Comparison

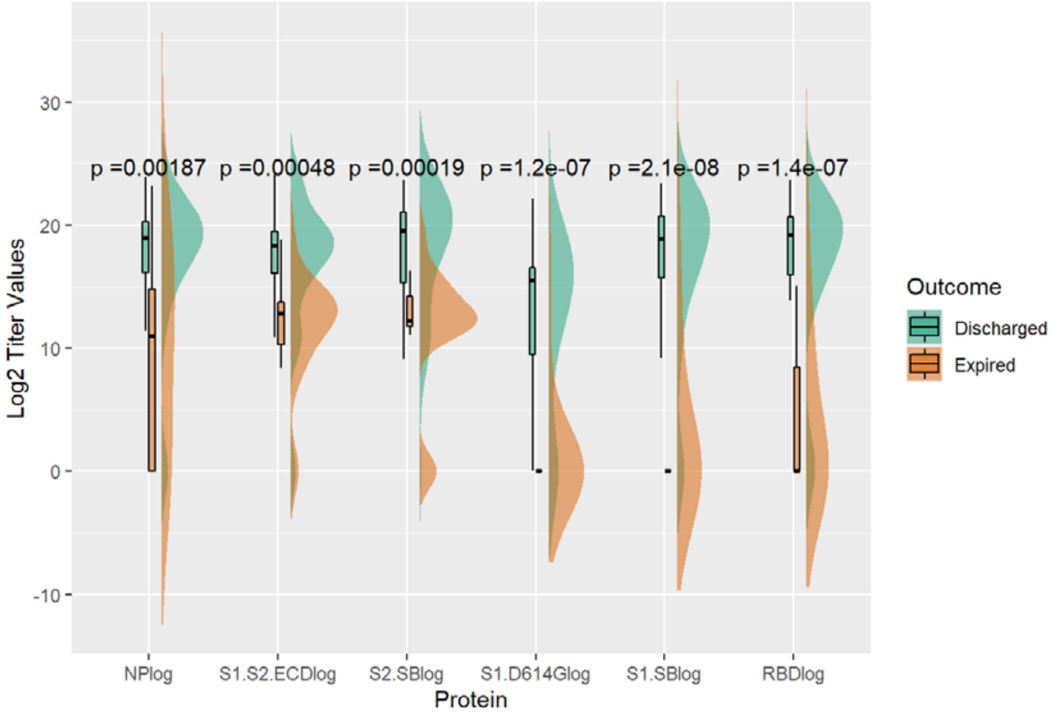


**Figure S16 IgG Cut-point MFI Week 1 Comparison (exclude 1000 and 1007).** Distribution of IgG antibody levels (log transformed) at week 1 post onset in expired and discharged groups with t-test comparison p-value.

### IgG Sensitivity Analysis Modeling

#### Table S5. Estimates from Linear Mixed Model (Model 1) of Difference between Discharged and Expired Groups in IgG Antibody Levels (log) by Time

| | Average Difference between Discharged and Expired groups | | | | | |
|---|---|---|---|---|---|---|
| | NP | S1 | S2 | S1S2ECD | S1D614 | RBD |
| Week1 | 2.91 | 8.68* | 4.16* | 3.33 | 6.50* | 6.28* |
| Week2 | 2.03 | 1.97 | 0.13 | 0.32 | 2.06 | 1.25 |
| Beyond Week2 | 1.78 | 2.21 | 1.54 | 1.37 | 1.03 | 2.05 |
| Change from week1 to week2 | -0.88 | -6.72* | -4.02* | -3.00* | -4.45* | -5.03* |
| Change from week1 to beyond week2 | -1.13 | -6.48* | -2.61* | -1.95 | -5.47* | -4.23* |

*Statistically Significant Differences in IgG Titer Values between Expired and Discharged groups at $\alpha = 0.05$

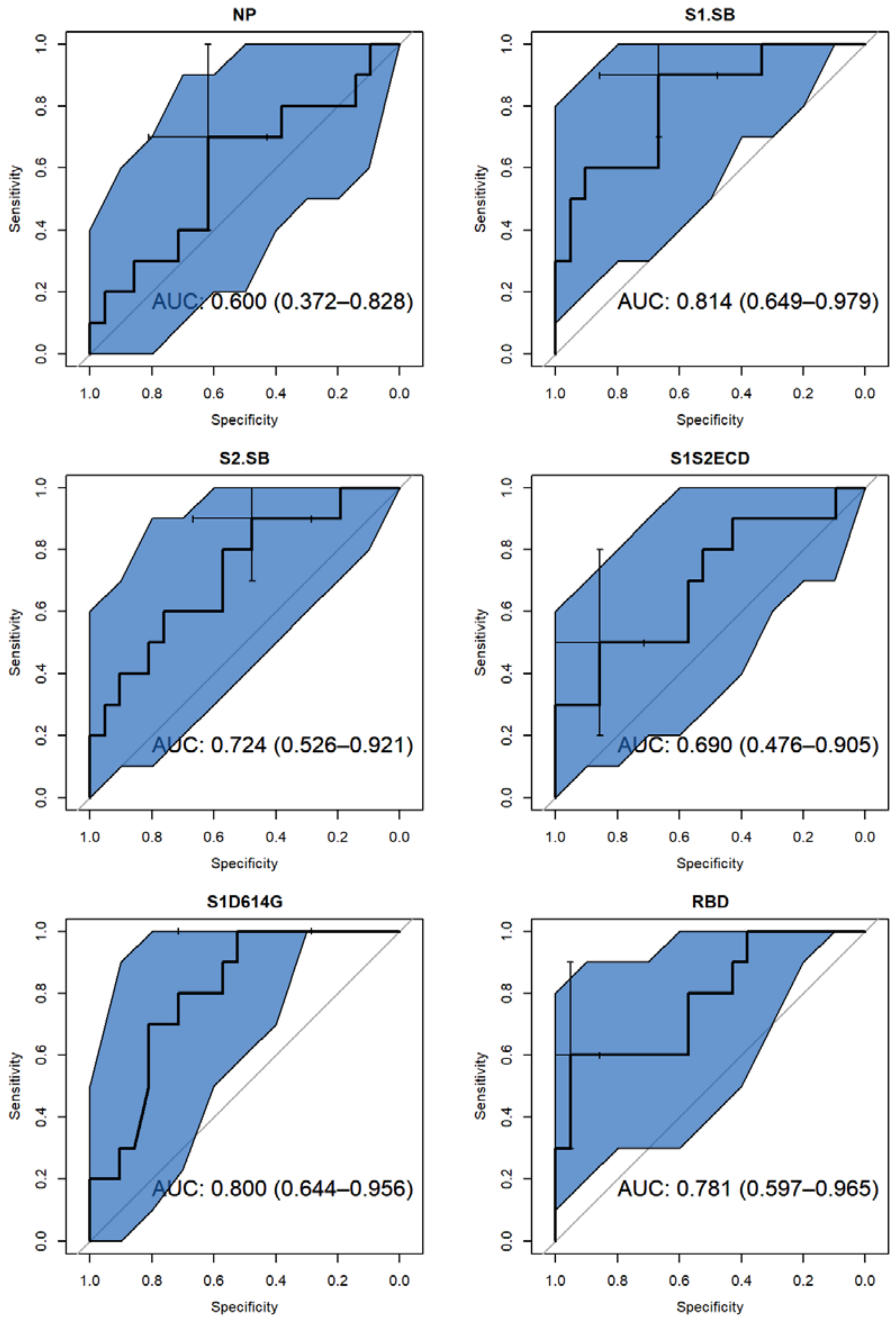


***Figure S17.*** *ROC curves for joint model (model 2) to predict death using IgG antibody levels; area under the curve (AUC) is presented within figure with 95% confidence intervals.*

### IgG Sensitivity Analysis (Males)

As in the expired group, only one patient out of 11 was female, to rule out any potential gender bias of our finding in discharged and expired groups, analyses were run for only male patients. Exclusion of female patients had no impact on the results.

### Anti-SARS CoV2 IgG Sensitivity Analysis Kinetics

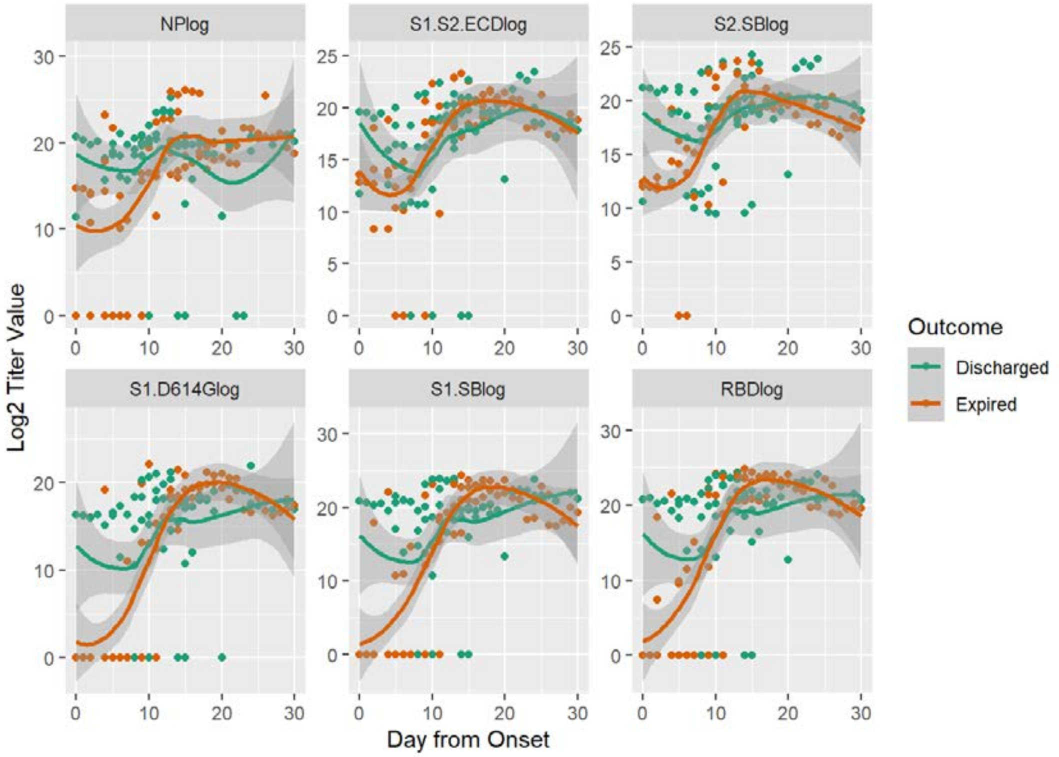


**Figure S18 Anti-SARS CoV2 IgG Kinetics (Males).** Observed IgG antibody titer values (log transformed) in expired and discharged groups of patients by protein antigen. Points are observed values for each patient at corresponding day from onset; lines are smoothed regression lines fit to the observed data with 95% confidence intervals.

### IgG Sensitivity Analysis MFI Week 1 Comparison

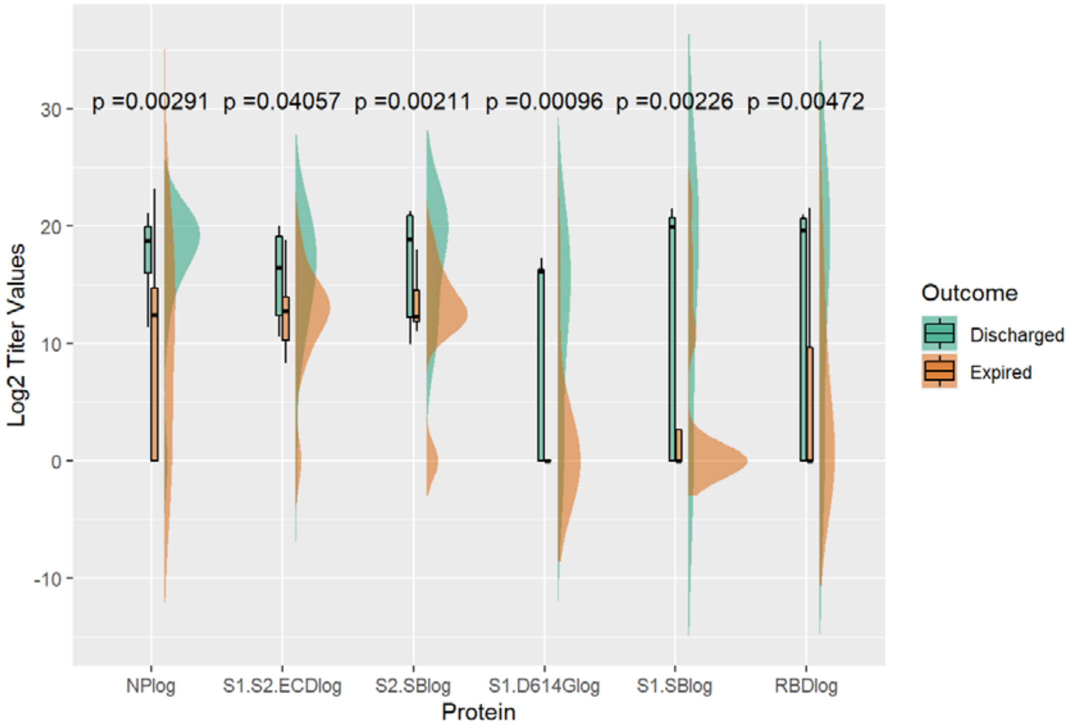


**Figure S19 IgG Cut-point MFI Week 1 Comparison (Males).** Distribution of IgG antibody levels (log transformed) at week 1 post onset in expired and discharged groups with t-test comparison p-value.

### IgG Sensitivity Analysis Modeling

#### Table S6. Estimates from Linear Mixed Model (Model 1) of Difference between Discharged and Expired Groups in IgG Antibody Levels (log) by Time

| | Average Difference between Discharged and Expired groups | | | | | |
|---|---|---|---|---|---|---|
| | NP | S1 | S2 | S1S2ECD | S1D614 | RBD |
| Week1 | 2.78 | 5.11 | 3.99* | 2.16 | 4.81 | 2.74 |
| Week2 | 1.27 | 0.96 | -0.22 | -1.46 | 1.04 | 0.45 |
| Beyond Week2 | 1.23 | 1.67 | 0.95 | 0.46 | 0.06 | 1.92 |
| Change from week1 to week2 | -1.51 | -4.14* | -4.21* | -3.62* | -3.77 | -2.29 |

| | | | | | | |
|---|---|---|---|---|---|---|
| Change from week1 to beyond week2 | -1.56 | -3.44 | -3.03* | -1.70 | -4.75* | -0.82 |
| *Statistically Significant Differences in IgG Titer Values between Expired and Discharged groups at $\alpha = 0.05$ | | | | | | |

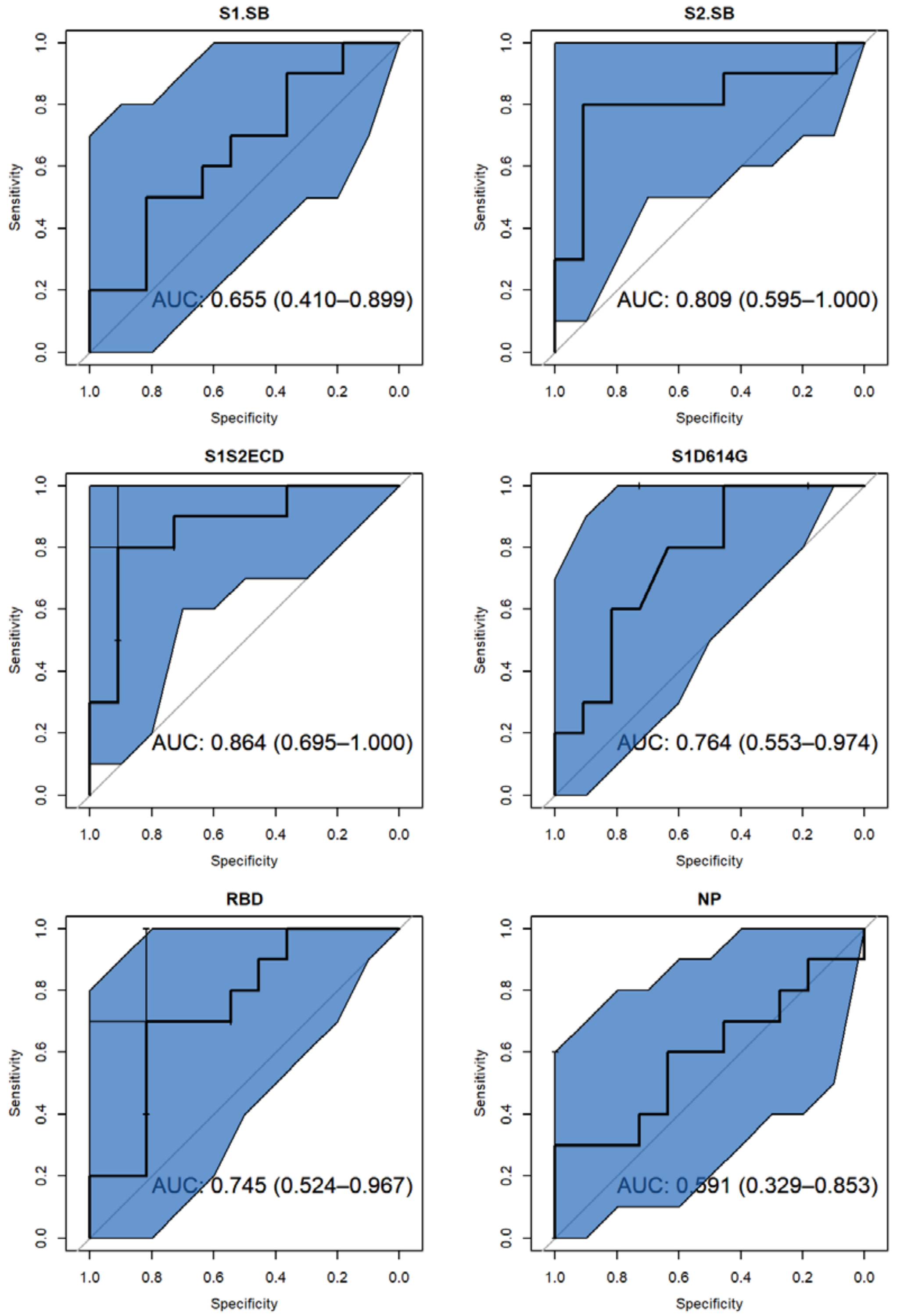

***Figure S20.*** *ROC curves for joint model (model 2) to predict death using Ig*G *antibody levels; area under the curve (AUC) is presented within figure with 95% confidence intervals.*
